# Long-read genome sequencing and variant reanalysis increase diagnostic yield in neurodevelopmental disorders

**DOI:** 10.1101/2024.03.22.24304633

**Authors:** Susan M. Hiatt, James M.J. Lawlor, Lori H. Handley, Donald R. Latner, Zachary T. Bonnstetter, Candice R. Finnila, Michelle L. Thompson, Lori Beth Boston, Melissa Williams, Ivan Rodriguez Nunez, Jerry Jenkins, Whitley V. Kelley, E. Martina Bebin, Michael A. Lopez, Anna C. E. Hurst, Bruce R. Korf, Jeremy Schmutz, Jane Grimwood, Gregory M. Cooper

## Abstract

Variant detection from long-read genome sequencing (lrGS) has proven to be considerably more accurate and comprehensive than variant detection from short-read genome sequencing (srGS). However, the rate at which lrGS can increase molecular diagnostic yield for rare disease is not yet precisely characterized. We performed lrGS using Pacific Biosciences “HiFi” technology on 96 short-read-negative probands with rare disease that were suspected to be genetic. We generated hg38-aligned variants and *de novo* phased genome assemblies, and subsequently annotated, filtered, and curated variants using clinical standards. New disease-relevant or potentially relevant genetic findings were identified in 16/96 (16.7%) probands, eight of which (8/96, 8.33%) harbored pathogenic or likely pathogenic variants. Newly identified variants were visible in both srGS and lrGS in nine probands (∼9.4%) and resulted from changes to interpretation mostly from recent gene-disease association discoveries. Seven cases included variants that were only interpretable in lrGS, including copy-number variants, an inversion, a mobile element insertion, two low-complexity repeat expansions, and a 1 bp deletion. While evidence for each of these variants is, in retrospect, visible in srGS, they were either: not called within srGS data, were represented by calls with incorrect sizes or structures, or failed quality-control and filtration. Thus, while reanalysis of older data clearly increases diagnostic yield, we find that lrGS allows for substantial additional yield (7/96, 7.3%) beyond srGS. We anticipate that as lrGS analysis improves, and as lrGS datasets grow allowing for better variant frequency annotation, the additional lrGS-only rare disease yield will grow over time.

## INTRODUCTION

Although genome and exome sequencing (GS/ES) are increasingly used to identify molecular causes of rare diseases, reported diagnostic rates range from 20-60% (Srivastava et al. 2019; Baxter et al. 2022), indicating that many conditions suspected to be genetic remain refractory to genomic testing. While some tested individuals may have phenotypes resulting from polygenic and/or environmental risk factors (e.g., Niemi et al. 2018), a subset of undiagnosed cases likely result from genetic factors that we are as-yet unable to identify. It is well-known that short-read genome sequencing (srGS) has poor sensitivity to many types of variants, especially structural variants (SVs) and variants affecting repetitive sequences (Wenger et al. 2019; Sanghvi et al. 2018; Mahmoud et al. 2024). Long-read genome sequencing (lrGS), in contrast, has been shown to greatly improve sensitivity to many of the variants missed by srGS (Logsdon et al. 2020), in addition to facilitating *de novo* assemblies to allow for more effective evaluation of structural variation (Cheng et al. 2021). Accordingly, lrGS has great potential to improve rare disease diagnostic testing and has been applied to several rare disease cohorts (Cohen et al. 2022; Miller et al. 2021; Hiatt et al. 2021).

In addition to changes in sequencing technology, the scope of knowledge about genes and our ability to annotate genetic variants has steadily increased. As such, systematic reanalysis of GS/ES data also leads to the discovery of previously overlooked clinically relevant variants, with diagnostic yield increases ranging from 4-31% depending on a variety of factors, most notably time since the previous analysis (Hiatt et al. 2018; Liu et al. 2019; Schobers et al. 2022; Hartley et al. 2023). While a variety of factors contribute to reanalysis discoveries, they often result from the discovery of new disease genes, which contributes to 42-75% of reanalysis findings (Hiatt et al. 2018; Liu et al. 2019; Schobers et al. 2022; Hartley et al. 2023). This reflects the rapid pace of discovery of new disease genes in the rare disease research community, which has been facilitated by data sharing via the MatchMaker Exchange and GeneMatcher (Philippakis et al. 2015; Sobreira et al. 2017).

Here we discuss findings from lrGS on a cohort of 96 short-read-negative cases, drawn from several studies focused on rare, suspected congenital diseases, especially early-onset neurodevelopmental disorders. We describe 19 relevant or potentially clinically relevant variants not previously evaluated or considered in 16 cases. We show that a combination of more comprehensive variant detection from lrGS and updated reanalysis contribute to these discoveries, supporting the value of a combination of lrGS and reanalysis as a strategy to maximize rates of discovery of highly penetrant variation leading to rare disease.

## RESULTS

We selected individuals with rare diseases who had undergone short-read exome sequencing (srES; n=2) or srGS (n=94) in previous research studies yet had no pathogenic or likely pathogenic variants (P/LP) nor variants of uncertain significance (VUS) identified (Bowling et al. 2017; East et al. 2021; Bowling et al. 2022). Most of our cohort consisted of children (89% were <18 years of age at time of enrollment) with a neurodevelopmental disorder (NDD, 70%), multiple congenital anomalies (MCA, 22%), or a suspected genetic myopathy (8%). Probands consisted of 66% males (63/96); genetically inferred ancestries for probands revealed 72% European (69/96), 21% African/African American (20/96), 3% Admixed American (3/96), 1% Southeast Asian (1/96) and 3% unspecified admixture ancestries (Table 1). For these 96 cases, we performed lrGS using Pacific Biosciences “HiFi” sequencing to a median depth of 27X (Table 1, Supplemental Table S1). For a subset (10/96), we also performed lrGS on parents (median parental HiFi depth of 22X, Supplemental Table S2). We also generated *de novo* assemblies for each proband using hifiasm (Cheng et al. 2021), with parental srGS used for kmer-based binning and phasing when available. The median N50 for all proband contigs was 29.05 Mb (Table 1).

**Table 1.** Demographics, and sequencing and assembly metrics for the cohort Sex.

|  |  |  |  |  |
| --- | --- | --- | --- | --- |
| **Sex** | Male | 63 (66%) |  |  |
|  | Female | 33 (34%) |  |  |
| **Predicted Major Genetic Ancestry** | European (EUR) | 71.88% |  |  |
|  | African (AFR) | 20.83% |  |  |
|  | Admixed American (AMR) | 3.13% |  |  |
|  | Southeast Asian (SAS) | 1.04% |  |  |
|  | Unspecified Admixed (UNKNOWN) | 3.13% |  |  |
| **Sequencing Metrics** |  | **Median** | **Min** | **Max** |
|  | Sequenced Bases (Gb) | 80.9 | 53.5 | 131.5 |
|  | Mean Read Length (Sequenced) | 16,771 | 11,295 | 21,412 |
|  | Median Coverage (X) | 27 | 18 | 44 |
|  | Percent Covered at 10x | 97.80% | 95.10% | 99.00% |
|  | Percent Covered at 20x | 84.90% | 36.10% | 97.50% |
| **Proband Assembly N50 (Mb)** | All contigs | 29.05 | 2.10 | 71.30 |
|  | Maternal contigs | 29.30 | 2.10 | 71.30 |
|  | Paternal contigs | 28.70 | 2.20 | 67.90 |
|  | hap1 contigs | 31.70 | 18.20 | 42.90 |
|  | hap2 contigs | 27.85 | 14.40 | 40.80 |
| **Small Variant Metrics (DeepVariant)** | SNVs | 4,404,564 | 3,997,350 | 5,299,670 |
|  | Total indels | 970,031 | 926,286 | 1,153,616 |
| **Structural Variant Metrics (pbsv)** | Total Structural Variants | 55,586 | 53,834 | 65,120 |
|  | Deletion | 24,757 | 23,799 | 29,531 |
|  | Duplication | 3,017 | 2,820 | 3,649 |
|  | Insertion | 27,594 | 26,441 | 32,437 |
|  | Inversion | 121 | 99 | 155 |
|  | Breakend | 193 | 124 | 312 |

HiFi reads were aligned to hg38, and variant calling was performed using DeepVariant (Poplin et al. 2018) and pbsv (https://github.com/PacificBiosciences/pbsv, see Methods). A median of 4.4 million SNVs and 970,031 indels were called in each proband (Table 1).

### Analysis of SVs

We detected a median of 55,586 SVs of varying classes across the 96 probands using pbsv (Table 1, Supplemental Table S3). We sought to characterize how filtering SVs by frequency could reduce manual curation burden by considering five allele frequency resources. First, we created a set of “cohort” SVs by performing SV call merging across all 96 probands using Jasmine (Kirsche et al. 2023) and generating allele counts from the merged set (set 1). We then used a second Jasmine merge step to match cohort SVs with SV frequencies from: an in-house set of 266 HiFi genomes including all cohort probands and parents, samples from other internal projects, and public HiFi data (set 2, see Methods); gnomAD v4 SV frequencies from 63,046 short read samples (set 3, Collins et al. 2020) Human Genome Structural Variant Consortium phase 2 (HGSVC2) assembly-based calls from 18 HiFi samples (set 4, Ebert et al. 2021); and a PacBio-provided set of pbsv calls from 103 HiFi samples from the Human Pangenome Reference Consortium (HPRC) and Genome in a Bottle (GIAB) consortia (set 5, Ebert et al. 2021; https://ftp.1000genomes.ebi.ac.uk/vol1/ftp/data_collections/HGSVC2/release/v2.0/integrated_callset/variants_freeze4_sv_insdel_alt.vcf.gz; https://github.com/PacificBiosciences/svpack/raw/main/resources/HPRC_GIAB.GRCh38.pbsv.vcf.gz). We note that these data are at least partially redundant with one another (e.g., set 1 is a partial subset of set 2 and many public samples such as NA12878 are shared between sets 2-5). However, our goal was to maximize filtering performance by including calls from as many datasets and discovery methods as possible. When filtering SVs with this data, we found probands had a median of 1,721 “rare” SVs, defined as having an allele frequency <1% in public SV databases (sets 3-5) and an allele count <4 in the cohort or internal (sets 1 or 2). A subset of these (proband median of 87 SVs) were within 50 bp of a RefSeq exon (Supplemental Table S3). We also filtered to identify “private” SVs as those absent from sets 3-5, having a set 1 allele count of 1, and having a set 2 allele count of <= 2 (allowing for parental inheritance). This filtering results in a median of 733 SVs per proband, only 40 of which are within 50 bp of a RefSeq exon (Supplemental Table S3).

### Findings from lrGS

lrGS SNVs/indels were annotated with features such as gene overlaps, coding consequences, computational impact scores, and allele frequencies. They were then filtered and analyzed using in-house software that is also used for srGS data (Hiatt et al. 2021). Rare SVs were assessed by visualization of reads in IGV and prioritization and analysis using SvAnna (Danis et al. 2022). All variants of interest were subject to curation using American College of Medical Genetics and Genomics and Association for Molecular Pathology (ACMG/AMP) and ClinGen criteria to identify potentially clinically relevant variation (Richards et al. 2015; Riggs et al. 2020). We identified 19 potentially “clinically relevant” variants, defined here as being pathogenic, likely pathogenic, or variants of uncertain significance (P/LP/VUS), in 16 of the 96 cases (Table 2). Seven of these have a case-level classification of Definitive Diagnostic or Likely Diagnostic, which we define as P/LP variants that likely fully explain the reason for testing (Bowling et al. 2022). The remaining nine cases have Uncertain case-level classifications, either due to the variants being VUSs or being P/LP variants in genes whose associated phenotypes do not closely match the observed phenotype. Findings in seven probands exemplify the unique benefits of lrGS and are highlighted below.

**Table 2.**
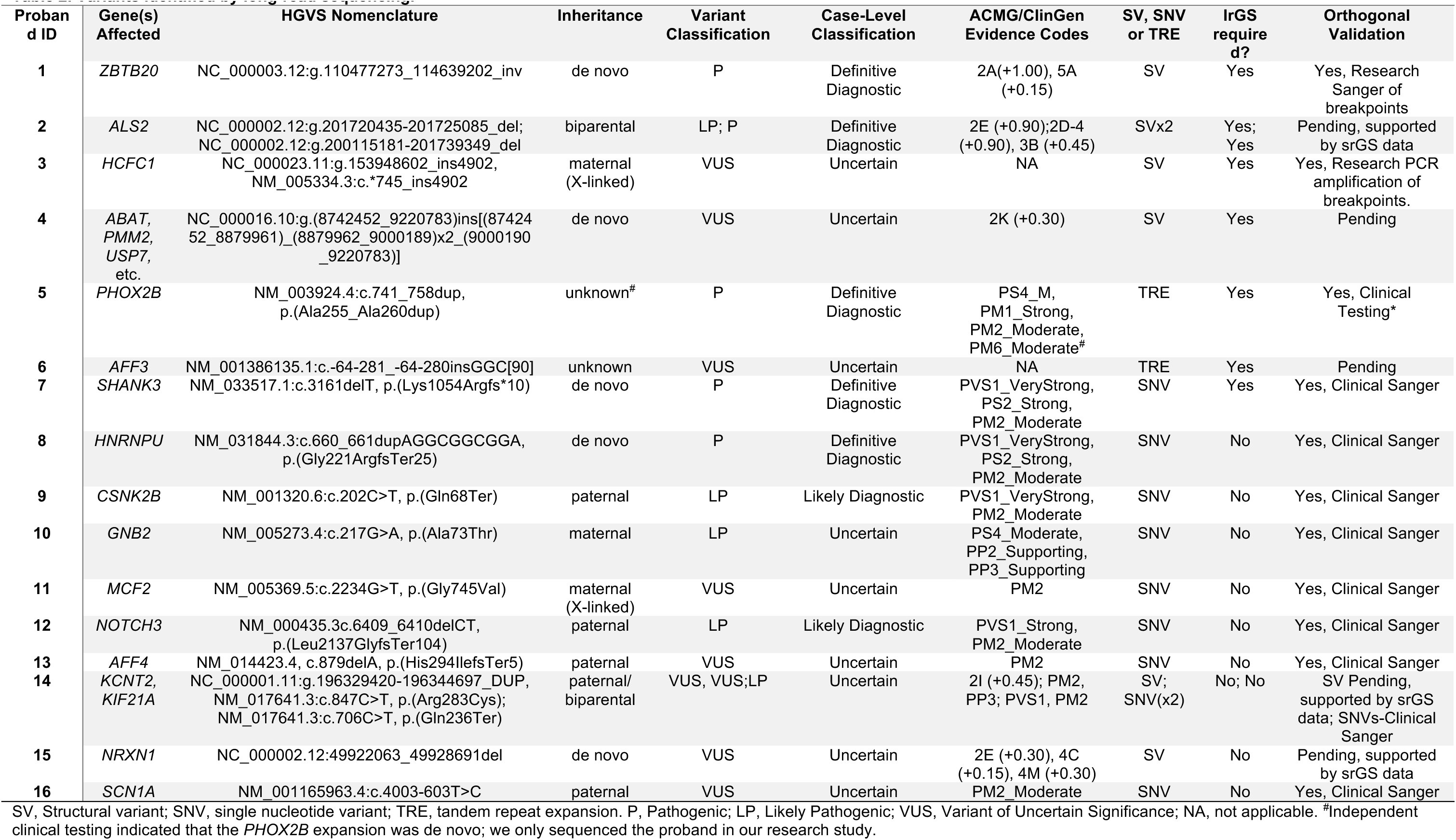
Variants identified by long-read sequencing.

| Proband ID | Gene(s) Affected | HGVS Nomenclature | Inheritance | Variant Classification | Case-Level Classification | ACMG/ClinGen Evidence Codes | SV, SNV or TRE | IrGS require d? | Orthogonal Validation |
| --- | --- | --- | --- | --- | --- | --- | --- | --- | --- |
| 1 | ZBTB20 | NC_000003.12:g.110477273_114639202_inv | de novo | P | Definitive Diagnostic | 2A(+1.00), 5A (+0.15) | SV | Yes | Yes, Research Sanger of breakpoints |
| 2 | ALS2 | NC_000002.12:g.201720435-201725085_del; NC_000002.12:g.200115181-201739349_del | biparental | LP; P | Definitive Diagnostic | 2E (+0.90);2D-4 (+0.90), 3B (+0.45) | SVx2 | Yes; Yes | Pending, supported by srGS data |
| 3 | HCFC1 | NC_000023.11:g.153948602_ins4902, NM_005334.3:c.*745_ins4902 | maternal (X-linked) | VUS | Uncertain | NA | SV | Yes | Yes, Research PCR amplification of breakpoints. |
| 4 | ABAT, PMM2, USP7, etc. | NC_000016.10:g.(8742452_9220783)ins[(8742452_8879961)_(8879962_9000189)x2_(9000190_9220783)] | de novo | VUS | Uncertain | 2K (+0.30) | SV | Yes | Pending |
| 5 | PHOX2B | NM_003924.4:c.741_758dup, p.(Ala255_Ala260dup) | unknown<sup>#</sup> | P | Definitive Diagnostic | PS4_M, PM1_Strong, PM2_Moderate, PM6_Moderate<sup>#</sup> | TRE | Yes | Yes, Clinical Testing* |
| 6 | AFF3 | NM_001386135.1:c.-64-281_-64-280insGGC[90] | unknown | VUS | Uncertain | NA | TRE | Yes | Pending |
| 7 | SHANK3 | NM_033517.1:c.3161delT, p.(Lys1054Argfs*10) | de novo | P | Definitive Diagnostic | PVS1_VeryStrong, PS2_Strong, PM2_Moderate | SNV | Yes | Yes, Clinical Sanger |
| 8 | HNRNPU | NM_031844.3:c.660_661dupAGGCGGCGGA, p.(Gly221ArgfsTer25) | de novo | P | Definitive Diagnostic | PVS1_VeryStrong, PS2_Strong, PM2_Moderate | SNV | No | Yes, Clinical Sanger |
| 9 | CSNK2B | NM_001320.6:c.202C>T, p.(Gln68Ter) | paternal | LP | Likely Diagnostic | PVS1_VeryStrong, PM2_Moderate | SNV | No | Yes, Clinical Sanger |
| 10 | GNB2 | NM_005273.4:c.217G>A, p.(Ala73Thr) | maternal | LP | Uncertain | PS4_Moderate, PP2_Supporting, PP3_Supporting | SNV | No | Yes, Clinical Sanger |
| 11 | MCF2 | NM_005369.5:c.2234G>T, p.(Gly745Val) | maternal (X-linked) | VUS | Uncertain | PM2 | SNV | No | Yes, Clinical Sanger |
| 12 | NOTCH3 | NM_000435.3c.6409_6410delCT, p.(Leu2137GlyfsTer104) | paternal | LP | Likely Diagnostic | PVS1_Strong, PM2_Moderate | SNV | No | Yes, Clinical Sanger |
| 13 | AFF4 | NM_014423.4, c.879delA, p.(His294IlefsTer5) | paternal | VUS | Uncertain | PM2 | SNV | No | Yes, Clinical Sanger |
| 14 | KCNT2, KIF21A | NC_000001.11:g.196329420-196344697_DUP, NM_017641.3:c.847C>T, p.(Arg283Cys); NM_017641.3:c.706C>T, p.(Gln236Ter) | paternal/ biparental | VUS, VUS;LP | Uncertain | 2I (+0.45); PM2, PP3; PVS1, PM2 | SV; SNV(x2) | No; No | SV Pending, supported by srGS data; SNVs-Clinical Sanger |
| 15 | NRXN1 | NC_000002.12:49922063_49928691del | de novo | VUS | Uncertain | 2E (+0.30), 4C (+0.15), 4M (+0.30) | SV | No | Pending, supported by srGS data |
| 16 | SCN1A | NM_001165963.4:c.4003-603T>C | paternal | VUS | Uncertain | PM2_Moderate | SNV | No | Yes, Clinical Sanger |

SV, Structural variant; SNV, single nucleotide variant; TRE, tandem repeat expansion. P, Pathogenic; LP, Likely Pathogenic; VUS, Variant of Uncertain Significance; NA, not applicable. <sup>#</sup>Independent clinical testing indicated that the *PHOX2B* expansion was de novo; we only sequenced the proband in our research study.

### lrGS-informed SVs

lrGS uncovered a *de novo*, 4 Mb, copy-neutral, paracentric inversion on chromosome 3 (NC_000003.12:g.110477273_114639202_inv) in Proband 1 (Figure 1). This inversion spans about 35 protein-coding genes, and one breakpoint of this inversion lies within an intron of *ZBTB20* (MIM: 606025). The event breakpoints are visible in both long and short reads for this proband, but it was only called as an inversion in lrGS. The variant is private to the proband and is predicted to disrupt *ZBTB20*. Loss-of-function (LOF) variation in *ZBTB20* is associated with Primrose Syndrome (MIM: 259050) and 3q13.31 Microdeletion Syndrome (Juven et al. 2020). Proband 1’s reported features include moderate intellectual disability (ID), delayed speech and language development, muscular hypotonia, strabismus, and hypoplastic corpus callosum. She is also non-ambulatory. Several of these features overlap Primrose Syndrome. We have classified this variant as pathogenic and the case-level designation is Definitive Diagnostic (Table 2).

**Figure 1.**
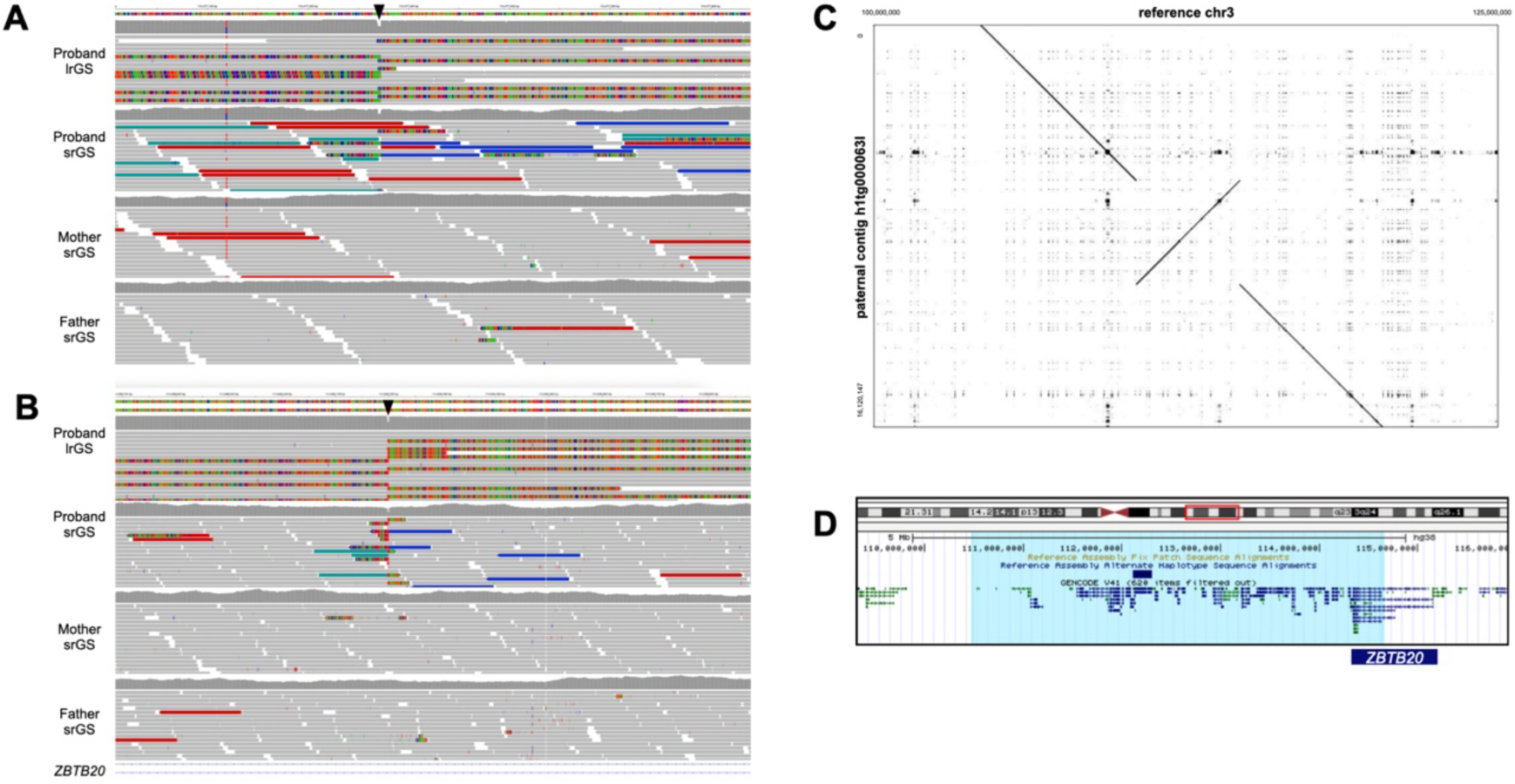
A *de novo, 4* Mb paracentric inversion in proband 1, affecting *ZBTB20.* A, B. Visualization of a subset of proband and parent reads in IGV at the 5’ (A) and 3’ (B) breakpoints (black arrowheads) indicate a de *novo* event. C. Alignment of the probanďs assembled paternal contig versus the reference genome supports the inversion. D. Visualization of the inverted region (highlighted in light blue) in the UCSC browser shows the inversion spans 35 protein-coding genes and likely disrupts the *ZBTB20* gene (dark blue bar).

Proband 2 presented with spasticity, ataxia, and leukodystrophy. srGS was negative, but lrGS identified two structural variants (SVs) identified in *trans* in *ALS2* (MIM: 606352). These include a maternally-inherited 4.65 kb deletion (chr2:201720435-201725085_del) that removes exons 21-23 of NM_020919.4, and a paternally-inherited ∼1.6 Mb deletion (chr2:200115181-201739349_del) that spans several genes, including the 3’ end of *ALS2* (deletion of exons 12-34 of NM_020919.4, Figure 2, Table 2). While these variants are visible in short-read data, the smaller deletion was called as a heterozygous deletion in the mother and as a homozygous deletion in the proband, obfuscating the nature of the variation and raising quality-control concerns. Given the results of the lrGS, the srGS variant calls logically resulted from the small maternal deletion intersecting with the larger, overlapping paternal deletion. This case highlights the difficulties in identification and analysis of overlapping SVs of unknown phase. Variation in *ALS2* is associated with several AR conditions (Juvenile amyotrophic lateral sclerosis 2, MIM: 205100; Juvenile Primary lateral sclerosis, MIM: 606353; and infantile onset ascending Spastic paralysis, MIM: 607225), each of which have features that overlap the proband’s presentation. These variants are classified as P/LP and, given the degree of overlap with expected phenotypes, the case-level designation is Definitive Diagnostic.

**Figure 2.**
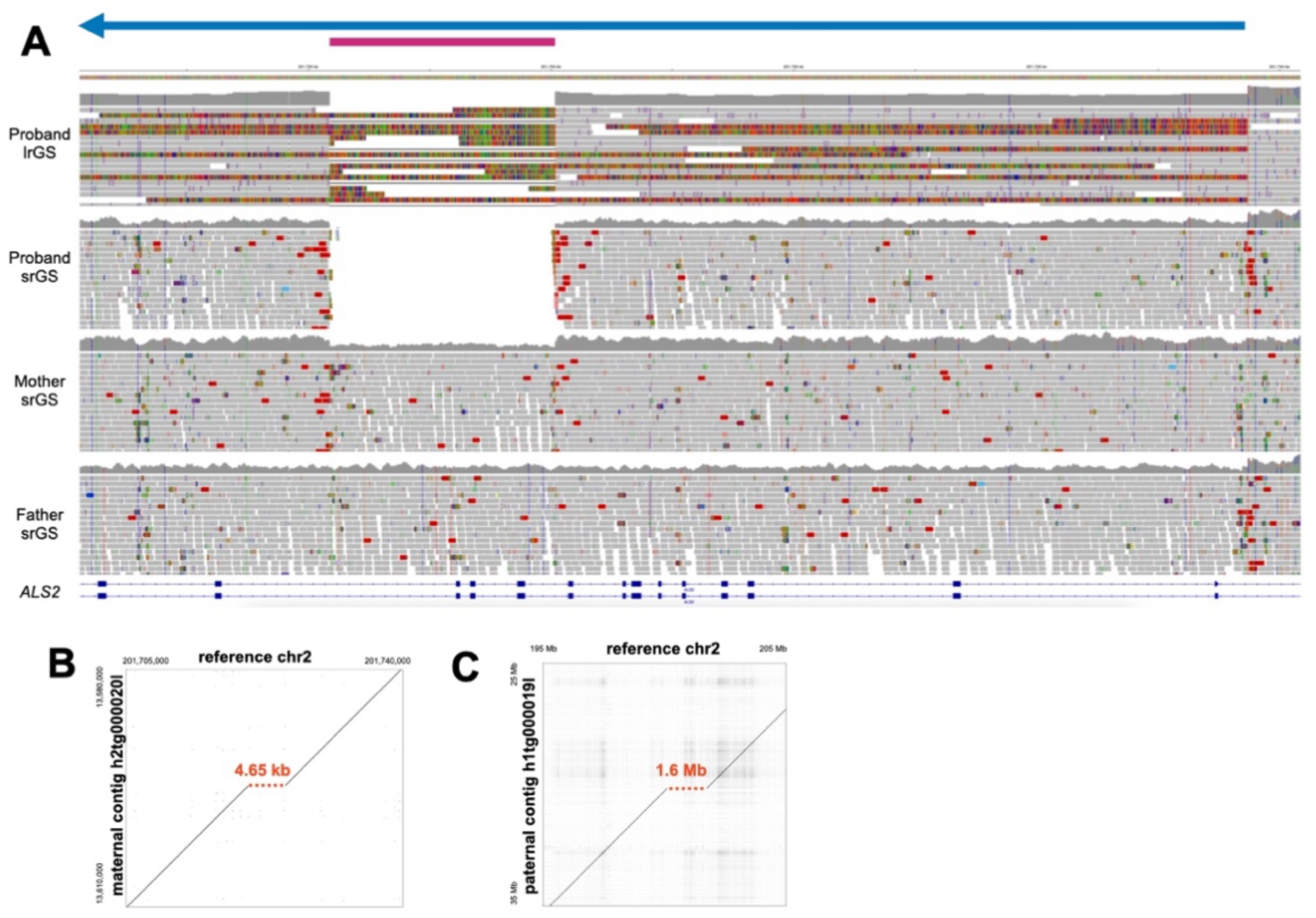
Two *ALS2* deletions in trans in Proband 2. **A.** Visualization of proband and parent reads in IGV indicate two overlapping deletions in *ALS2*; a smaller maternal deletion (pink bar) and a larger paternal deletion (blue bar/arrow). Alignment of the proband’s assembled maternal (B) or paternal contig (C) versus the reference genome support the two deletions (red dashed lines).

Proband 3 is a male with a strong X-linked family history of ID (Figure 3A). srES and srGS were both negative, although in srES, two neighboring SNVs were called, and manually curated, in the 3’ UTR of *HCFC1* (MIM: 300019). While srGS resulted in no calls in this region, visualization of reads in IGV suggested an insertion of unknown length and consequence (Figure 3C). Variation (mostly missense and proposed regulatory variation) in *HCFC1* has been associated with X-linked recessive Methylmalonic aciduria and homocysteinemia, cblX type (MIM: 309541). However, most affected individuals present with severely delayed psychomotor development, seizures, and methylmalonic aciduria. Proband 3’s family reported neither of the latter two features. Given the uncertainties in the identity, structure and consequence of the variants in both srES and srGS and the lack of clear phenotypic relevance for the gene, these variant calls were curated but not originally considered to be good candidates for clinical relevance. HiFi sequencing identified a 4,902 bp mobile element insertion (MEI) in the 3’ UTR of *HCFC1* (chrX:153948602_ins4902), consisting of both SVA and L1 sequence; this variant was likely inherited from a heterozygous carrier mother, as indicated by srGS reads at the breakpoints (Figure 3C). While this insertion does not affect protein-coding sequence, it is predicted to increase the length of the 3’ UTR by 3.75x (from ∼1800 nt to about ∼6800 nt). We subsequently performed 3’-end RNA-seq on blood from both the proband and his father, generating ∼8×10^6^ reads for each sample (see Methods). *HCFC1* shows the greatest expression decrease in the proband relative to his father, an ∼8.8-fold reduction, among all genes with at least 10 counts in each sample (Supplemental Figure S1). While these results are consistent with the hypothesis that the insertion has a large effect on *HCFC1* expression and potential activity, they are not definitive. Further, expression or segregation analyses in additional family members could not be assessed. Given the uncertainty of the molecular consequence, the differences in phenotypic features, and the lack of additional segregation data, we classified this as a VUS, with a case-level designation of Uncertain.

**Figure 3.**
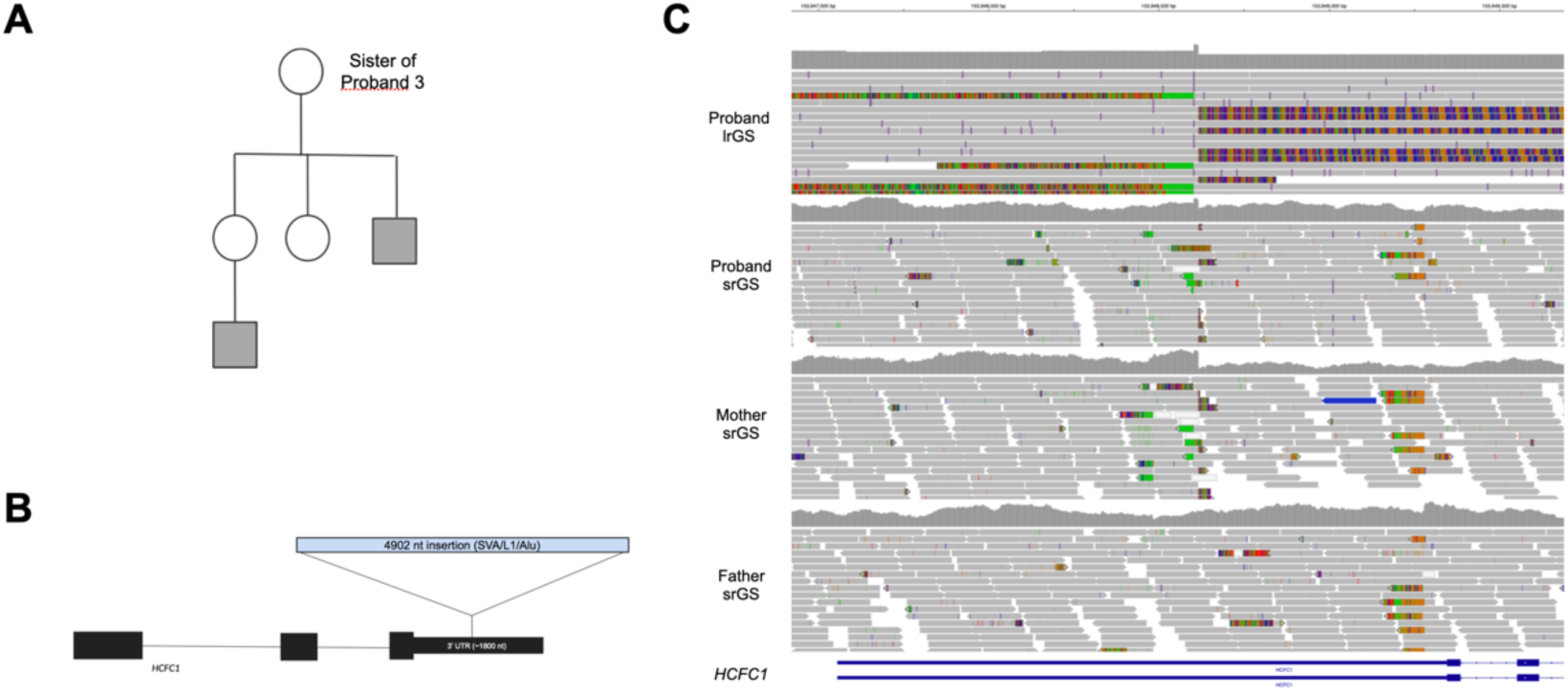
Proband 3 has a 4 kb insertion in the 3’ UTR of HCFC1. **A.** The probanďs family has a history of X-linked intellectual disability, as the proband (not shown) and two other male relatives (gray squares) are affected. B. Model of the relative length of the insertion in the 3’ UTR. C. The insertion is likely inherited from a heterozygous carrier mother, as indicated by srGS reads.

Proband 4 has a complex tandem *de novo* duplication affecting 16p13.2 (NC_000016.9:g.(8742452_9220783)ins[(8742452_8879961)_(8879962_9000189)x2_(9000190_9220783)], Table 2, Supplemental Figure S2). Overlapping duplications have been reported in gnomAD but are rare (Lek et al. 2016). One individual in Decipher (Patient: 251349) has also been reported with a very similar duplication of uncertain consequence (Deciphering Developmental Disorders 2015). These duplications span seven genes, three of which are associated with disease: *ABAT*, *PMM2*, and *USP7*. The first (*ABAT*) is intersected by a duplication breakpoint, but all other breakpoints lie within intergenic regions. *USP7* is the only gene associated with autosomal dominant disease (Hao-Fountain Syndrome, MIM:616863), but this gene is expected to have a copy number of four in this proband, while LOF is generally the mechanism associated with disease (Hao et al. 2015). Some general features of Hao-Fountain Syndrome overlap this proband, but it is not a strong phenotypic fit. The proband is reported to have moderate ID, seizures, microcephaly, and facial dysmorphisms. Proband 4 first had a trio srES and no variants were returned, and this duplication was not called. Trio lrGS identified the order, orientation, and copy number of segments of these tandem duplications. We have classified this variant as a VUS, with a case-level designation of Uncertain. Note that this variant is pending orthogonal validation. Proband 4 was one of the two individuals who only previously had srES rather than srGS, and this duplication was not easily visualized in srES data.

### lrGS-informed SNV/Indels

Improved variant calling in repeat regions is also a benefit of lrGS (Nurk et al. 2022). In addition to analysis of SNVs and SVs in our standard pipeline, we assessed variant calls in 66 tandem repeat expansion (TRE) regions, including both known disease-associated and candidate disease-associated loci (Supplemental Table S4). We intersected TRE regions of interest with pbsv-called variants in each individual and compared these calls to known pathogenic expansion sizes from the literature. We identified several large heterozygous insertions in repeat regions that, while longer than the expected pathogenicity threshold, were predicted after manual curation to be benign based on their sequence content (Nakamura et al. 2020, Supplemental Figure S3A). We also identified two large heterozygous insertions in *RFC1* (MIM: 102579), one of which is benign based on sequence content and one of which is expected to be a pathogenic insertion. However, *RFC1*-associated disease (CANVAS, MIM: 614575) is caused by biallelic expansions, which we did not observe (Supplemental Figure S3B), suggesting this proband is merely a heterozygous carrier.

In Proband 5, we observed a *de novo* 18-bp alanine tract expansion in *PHOX2B* (MIM: 603851, NM_003924.4:c.741_758dup, p.(Ala255_Ala260dup), Table 2, Supplemental Figure S4), associated with Central Hypoventilation Syndrome, with or without Hirschsprung Disease (MIM: 209880). This disorder was clinically suspected, but variation in *PHOX2B* was missed by initial clinical genetic testing and srGS, which was performed on a PCR+ srGS library. This variant is reported as pathogenic in literature (Amiel et al. 2003), is a strong match to the proband’s observed symptoms, and was confirmed independently by additional clinical testing. We have classified this variant as pathogenic, with a case-level classification of Definitive Diagnostic.

In proband 6, we identified a 270 bp insertion in *AFF3* (MIM:601464, NM_001386135.1:c.-64-281_-64-280insGGC[90], Table 2, Supplemental Figure S5). While missense variants in *AFF3* have been associated with KINSSHIP Syndrome (MIM:619297), an expansion of a CGG-repeat in the promoter of this gene and subsequent hypermethylation of the promoter has recently been reported to be associated with NDDs (Jadhav et al. 2023). The majority of probands in our cohort (77/96) had both alleles matching hg38 in this region. Among the 19 probands harboring non-reference alleles, proband 6 had a 270 bp insertion, and the remaining 18 probands had insertions ranging from 24-84 bp in length (8-28 triplet repeat units). Similarly, when comparing to the larger, “set 2” database of HiFi genomes (n=266), only 53/266 individuals harbor a non-reference allele; the longest insertions outside of Proband 6 are 105 bp (35 repeats) and 93 bp (31 repeats), each in different individuals, while the median non-reference insertion is 36 bp (12 repeats). Thus, the 270 bp insertion (90 repeat units) in Proband 6 is a clear outlier, being nearly three-fold longer than the second longest insertion (Supplemental Table S4, Supplemental Figure S6). Proband 6 was sequenced as a neonate and presented with intrauterine growth restriction (IUGR) and hypoplastic left heart (HLH). Jadhav et al. suggest that normal variation ranges to up to ∼38 repeat units, with >= 61 repeats being a likely pathogenic threshold; our results are consistent with those observations. Nevertheless, there is a need to further replicate and confirm the spectrum of normal and pathogenic variation in *AFF3* repeat lengths. Given this uncertainty and the uncertainty regarding the proband’s cognitive development, we have classified this TRE as a VUS, and the case-level classification is Uncertain. Also note that this variant is pending orthogonal validation.

A *de novo SHANK3* SNV (NM_033517.1:c.3161delT, p.(Leu1054Argfs*10)) was identified by lrGS in proband 7. While two reads in the srGS data support this 1 bp deletion, the variant was not called by our srGS variant calling pipeline (Supplemental Figure S7). LOF variation in *SHANK3* (MIM: 606230) is associated with Phelan-McDermid syndrome (MIM: 606232). Features of this syndrome are consistent with the proband’s features, and we classified this variant as pathogenic (case-level Definitive Diagnostic). Coverage of this region in short read data does not indicate a systematic coverage deficiency, as mean coverage within 50 bp of this variant in the srGS data for the cohort is 26.1x (n=96), while it is 15.1x for proband 7. Further, this gene is not present in the list of medically relevant genes that tend to be poorly covered by srGS (Wagner et al. 2022), suggesting it may have resulted from a stochastic loss of alternative allele reads in the srGS data. Nevertheless, lrGS has been shown to provide better overall sensitivity and specificity to SNVs and indels in Genome-In-A-Bottle (GIAB) gold-standard datasets compared to srGS (Logsdon et al. 2020; Hiatt et al. 2021) and thus detection failures to variants such as this *SHANK3* event are more likely in srGS data in general.

### Reinterpretation of SNVs

The remaining 9 cases that had relevant variation identified following lrGS illustrate the value of reanalysis and/or reinterpretation (Table 2, Supplemental Case Reports). In three cases (Probands 8-11, *HNRNPU, CSNK2B, GNB2, MCF2*) we identified variation in genes that had additional published support for association of the gene or the variant with disease since the time of the most recent analysis. In another four cases (Probands 12-15, *NOTCH3, AFF4, KCNT2, KIF21A, NRXN1*) we identified variation in established disease genes that conflicted with the published data regarding molecular mechanisms or expected mode of inheritance. For these cases, the SNVs were also identified by srGS but they were prioritized more effectively following lrGS, particularly after no new P/LP/VUS SVs of interest were observed. We note that in 7/9 cases, variants were identified in an unaffected or mildly affected parent, which was somewhat unexpected in these cases due to suspicion of high penetrance (also see Supplemental Case Reports). Lastly, we identified a variant of interest in *SCN1A* that resulted from a targeted analysis of candidate “poison exon” variants (Proband 16, Felker et al. 2023).

## Discussion

There remains a considerably large fraction of disease suspected to have genetic causes that is refractory to genomic testing, a finding that has been repeatedly shown across many clinical and research projects (e.g., Srivastava et al. 2019; Baxter et al. 2022). Several non-mutually exclusive hypotheses exist to explain these observations. Environmental risk factors, such as teratogenic exposure or infectious disease, may be relevant to some phenotypes. Multigenic contributors are also likely to explain at least some cases. For example, a small but appreciable fraction of “double diagnoses”, in which an affected individual is observed to harbor two distinct diseases resulting from highly penetrant variation in two distinct genes, has been observed in clinical genomic studies (e.g. Posey et al. 2017). Notably, such discoveries are typically only made when the variation in both genes is independently amenable to a pathogenicity determination (e.g., would be P/LP regardless of P/LP variation in the other gene), and it is possible if not probable that at least some conditions result from combinations of variants in different genes that are not pathogenic in isolation (Papadimitriou et al. 2019). At the further end of this spectrum is a polygenic accumulation of many risk-factor alleles, which is known to be relevant to many common, complex diseases and which may contribute to some rare conditions, as has been suggested for at least a subset of neurodevelopmental disorders (Niemi et al. 2018).

We find it likely that a substantial fraction of unexplained rare disease arises from highly penetrant, monogenic variation that we have not yet been able to precisely identify or confidently interpret. The results from this study are consistent with that hypothesis, with over 15% of probands with previously negative testing being now found to harbor a relevant or potentially relevant genetic variant. Further, these observations are consistent with the general picture of rare disease testing in recent years. With the advent of exome capture and sequencing ∼15 years ago (Ng et al. 2010, 2009) and subsequent improvements in cost and efficiency, genome-wide detection of highly penetrant variation has greatly accelerated in recent years (Bamshad et al. 2019; Baxter et al. 2022; Hamosh et al. 2022; Boycott et al. 2022). Long-read genome sequencing represents the next phase of that acceleration by facilitating a substantial increase in variant comprehensiveness and accuracy. We note that this improvement derives from both sensitivity and specificity improvements. For example, effectively all the variation we describe in this study as being newly visible within lrGS is, in fact, visible in srGS data, at least at the breakpoint levels. However, being retrospectively visible, once the location and structure of a variant is known to exist, is a much lower bar than the ability to prospectively detect, define, filter, and curate such variation. For example, we describe here a 4.9 kb insertion of SVA and L1 sequence into the 3’ UTR of *HCFC1*. In retrospective analysis, the Mobile-Element Locator Tool (MELT, Gardner et al. 2017) detected a 1.2 kb SVA mobile element at this location in srGS. However, this call is incorrect with respect to size and sequence composition and MELT generally produces too many calls in our srGS data to allow for a sustainable level of manual curation (Hiatt et al. 2021). For example, this proband had 1,311 total MELT calls, 203 of which are within 50 bp of a RefSeq exon. Further, this individual call has an unbalanced number of reads supporting the left and right breakpoints for this event (LP=17; RP=1). The low number of reads supporting the right breakpoint and the incorrect length of the insertion appear to be due to the presence of L1 sequence at the 3’ end of the insertion, compounded with a long polyA sequence (79 nt).

Our results are also consistent with other studies of the benefits of lrGS for discovering genetic contributors to disease. In 2021, for example, we showed in a small pilot project that 2 of 6 previously srGS-negative probands harbored clinically relevant variation uniquely interpretable by lrGS (Hiatt et al. 2021). Since that time, several other studies have also used lrGS for molecular diagnosis of rare disease. For example, Cohen and colleagues showed increased yield of GS (both lrGS and srGS) in exome-negative cases (Cohen et al. 2022). Unique discoveries for lrGS included detection of novel repeat expansions of *STARD7* and a compound heterozygous SNV/deletion that was easier to detect in lrGS. However, the increase in diagnostic yield (∼13%) was mainly from SVs detectable by either lrGS or srGS. Other studies have also shown the diagnostic value of lrGS, especially in small, well-phenotyped cohorts or families (Sakamoto et al. 2024; Kilich et al. 2024; Audet et al. 2023; Del Gobbo et al. 2023; Fukuda et al. 2023; Miller et al. 2021). Our results are similar to these studies at a high-level. However, some important differences are worth noting. One notable difference is that we have shown the value of lrGS in singletons. While srGS data was available for most of the probands’ parents, only 10/96 probands had parent lrGS data. Our filtering strategies were sufficient to allow prioritization of proband variants without parental data; further, once flagged for interest, inheritance data could often be assessed by looking at parental srGS reads. Based on this experience, prioritization of proband sequencing with targeted validation in parents is an effective way to increase diagnostic yield with lrGS while reducing lrGS sequencing needs.

We have also provided a direct, systematic comparison of lrGS to contemporaneously analyzed srGS in previously negative cases. Our results thus allow for the separation and description of clinically relevant variants that are “new” by virtue of being truly unique to lrGS versus those that are “new” by virtue of reanalysis, and which could be found via lrGS or srGS-reanalysis. In that context, we note that the benefits of reanalysis of older srGS data remain considerable. In the results described here, the “new” discoveries in 9 of 16 cases in lrGS were called correctly and interpretable within srGS data. These observations reflect the rapid pace of gene discovery in rare disease (Baxter et al. 2022; Boycott et al. 2022; Hamosh et al. 2022), and are consistent with other studies. For example, we previously showed that the probability of a negative srGS dataset harboring a clinically relevant variant increased from 1% within one year of a previous analysis to ∼22% if more than three years have passed since a previous analysis (Hiatt et al. 2018). Several other studies found similar results, with many reanalysis findings being due to recent publications of new gene-disease associations (Liu et al. 2019; Schobers et al. 2022; Hartley et al. 2023).

One possible optimal path to maximizing overall yield in previously srGS-negative individuals is to include reanalysis prior to lrGS. However, this reflects a cost/benefit ratio that depends on the cost of the analysis step in relation to the costs of sequencing. As lrGS costs decrease there may reach a point where the cost of variant analysis alone (which requires both compute resources and manual curation time and thus represents a non-trivial cost) is substantial relative to lrGS costs per se and which might favor a process of simply performing lrGS. Further, lrGS allows analysis of a more complete genomic picture, including both SNVs and larger variant types that are truly not called in srGS data. This can allow more accurate and confident variant prioritization (including SNVs) as evidenced by several of our cases.

We note that the results in this study were generated over a period of time with considerable change in lrGS protocols. For example, most probands in this study were sequenced on Sequel IIE machines (n=64) before the Revio (n=32) became available. Given the costs of data production, the 64 Sequel IIE samples were covered at lower-depth (median coverage 24.65X) than the 32 Revio-sequenced samples (median coverage 30.09X, Supplemental Figure S8), which may have reduced our sensitivity to variants in some loci in the earlier samples. Further, methylation data were not available in the early period of this study, although methylation calls are now routine and available for the most recent 44 probands, which may also impact diagnostic yield. For example, evaluation of the, at present VUS, *AFF3* expansion (Supplemental Figure S5, S6) would benefit from an assessment of methylation levels at this locus, as hypermethylation is likely associated with disease risk (Jadhav et al. 2023). Additionally, some probands harbor methylation alterations that are clinically relevant even in the absence of a pathogenic genetic variant (Aref-Eshghi et al. 2021).

In addition to changes in sequencing, informatic changes have also been considerable over the course of the data generation for this study. While we present and describe a uniform set of variant-calls, assemblies, and annotations (see Methods), analysis of individual samples took place simultaneously with the optimization of variant-calling and annotation pipelines. One particularly relevant change is variant-frequency annotations. While the key strength of lrGS is its ability to see variants that are invisible or poorly detected in srGS, the ability to discriminate genuine highly penetrant variation from the background of benign alleles depends on the ability to annotate and remove alleles that are common in the general population. As the main allele frequency resources are built from srGS data (e.g., Lek et al. 2016), we have limited ability to filter away likely benign alleles among the variants uniquely detected by lrGS. This ability, however, improved as more lrGS datasets were produced over the course of this study (Ebert et al. 2021; Nurk et al. 2022). Projects like the COLORs consortium (https://colorsdb.org/) are likely to improve frequency annotation in the future and continue to improve variant curation efficacy. Accumulating lrGS data from as many samples and studies as possible is critical for the long-term maximization of lrGS benefits.

In sum, rare disease genetics continues to be a rapidly advancing field. With data-sharing (Sobreira et al. 2015; Philippakis et al. 2015) and technology improvement (Wenger et al. 2019), the overall diagnostic yield for individuals with rare disease is increasing at a considerable pace each year. In that context, lrGS is a decisive improvement over srGS, providing substantial gains to variant specificity and sensitivity, especially for complex and repeat-associated variants. We anticipate that the degree of improvement will widen over time, as sequencing and analysis pipelines mature and as lrGS datasets grow.

## METHODS

### Short-read sequencing and variant calling

Probands, their parents, and, when appropriate, affected siblings were enrolled in one of four research studies aimed at identifying genetic causes of rare disease (Bowling et al. 2017; East et al. 2021; Bowling et al. 2022 and Pediatric Genomics (PGEN), unpublished). These studies were monitored by Western IRB and UAB IRB (WIRB 0071, UAB IRB protocols 170303004, 300000328, and 130201001). A parent or legal guardian gave consent for the proband, and assent was also obtained from those probands who were capable. In some cases, adult probands who were capable consented to participation in the study. All enrolled individuals consented to publication of de-identified data. Short read exome (srES) or short-read genome sequencing (srGS) was performed as described (Bowling et al. 2022; East et al. 2021; Hiatt et al. 2021; Bowling et al. 2017). Briefly, whole blood genomic DNA was isolated using the QIAsymphony (Qiagen), and sequencing libraries were constructed by the HudsonAlpha Genomic Services Lab or the Clinical Services Laboratory, LLC, using a standard protocol that generally included PCR amplification (86/96). Genomes were sequenced at an approximate mean depth of 30X, with at least 80% of base positions reaching 20X coverage. Exomes were sequenced to a mean depth of 71X. For short read reanalysis, srES and srGS reads were aligned and small variants and CNVs were called with a uniform pipeline to hg38, and SNVs/Indels and CNVs were curated using an in-house software tool, as previously described (Hiatt et al. 2021). Expected sample relatedness was confirmed with Somalier (v. 0.2.10, (Pedersen et al. 2020) and predicted major genetic ancestries were calculated with peddy (v. 0.4.1, Pedersen and Quinlan 2017).

### Long-Read sequencing, variant calling, analysis and de novo assemblies

Long-read sequencing was performed using HiFi (CCS) mode on either a PacBio Sequel II or Revio instrument (Pacific Biosciences of California, Inc.). Libraries were constructed using a SMRTbell Template Prep Kit (V1.0, 2.0 or 3.0) and tightly sized on a SageELF or BluePippin instrument (Sage Science, Beverly, MA, USA). Sequencing was performed using a 2 hour pre-extension with either 24 or 30 hour movie times. The resulting raw data was processed using either the CCS3.4 or CCS4 algorithm, as the latter was released during the course of the study. Comparison of the number of high-quality indel events in a read versus the number of passes confirmed that these algorithms produced comparable results. Probands were sequenced on 2-3 Sequel II or one Revio SMRT cell. Top off sequencing was performed if the sequencing did not meet the desired coverage (>20X). This resulted in an average estimated HiFi read depth of 26.1X (range 16.7-41) of raw, unaligned sequence data for probands. For 10 families, parents were also sequenced on 2-3 Sequel II SMRT cells, with an average estimated depth of 21.6x (range 14-28) of raw, unaligned sequence data for parents. Aligned sequencing metrics are shown in Supplemental Tables S1 and S2. HiFi reads were aligned to the were aligned to the hg38 no-alt analysis set (https://ftp.ncbi.nlm.nih.gov/genomes/all/GCA/000/001/405/GCA_000001405.15_GRCh38/seqs_for_alignment_pipelines.ucsc_ids/GCA_000001405.15_GRCh38_no_alt_analysis_set.fna.gz) using pbmm2 (v. 1.10.0). SNVs and small indels,were called with DeepVariant (v. 1.5.0) and used to haplotag the aligned reads with whatshap (v. 1.7). Structural variants were called using pbsv v2.9.0 (https://github.com/PacificBiosciences/pbsv). For the 10 cases with parent lrGS, candidate de novo SVs required a proband genotype of 0/1 and parent genotypes of 0/0, with ≥ 6 alternate reads in the proband and 0 alternate reads, and ≥5 reference reads in the parents.

De novo assemblies were generated for all probands using hifiasm (v. 0.15.2 (r334 or v.0.16.1-r375, Cheng et al. 2021). Hifiasm was used to create two assemblies. First, the default parameters were used, followed by two rounds of Racon (v1.4.10) polishing of contigs. For cases with parent GS data, trio-binned assemblies were built using kmers (srGS). The kmers were generated using yak (v0.1) using the suggested parameters for running a hifiasm trio assembly (kmer size=31 and Bloom filter size of 2**37). Maternal and paternal contigs went through two rounds of Racon (v1.4.10) polishing. Individual parent assemblies were also built with hifiasm (v0.15.2) using default parameters. The resulting contigs went through two rounds of Racon (v1.4.10) polishing.

Coordinates of breakpoints were defined by a combination of assembly-assembly alignments using minimap2 (Li 2018) (followed by use of bedtools bamToBed), visual inspection of CCS read alignments, and BLAT. Dot plots illustrating sequence differences were created using Gepard (Krumsiek et al. 2007).

### Structural Variant Merging and Filtering

Overall structural variant counts in Supplemental Table S1 were generated from the proband pbsv VCFs using bcftools (v1.15.1) and awk (e.g., bcftools filter -i ‘SVTYPE==”DEL” <input.vcf> | {a[i++]=$1; sum+=$1} END{asort(a); min=a[1]; max=a[i]; if(i%2==1) median=a[int(i/2)+1]; else median=(a[i/2]+a[i/2+1])/2; mean=sum/i; print mean, median, min, max;).

An internal allele frequency callset was constructed and periodically updated using all available internally sequenced HiFi pbsv variant calls and pbsv calls generated from public HiFi sequencing data (n=266 at the end of this analysis). Samples included a majority of participants from this cohort and our previous NDD pilot cohort (Hiatt et al. 2021, 120/266); samples from HudsonAlpha non-NDD projects (35/266); HudsonAlpha-sequenced data from HG001, HG003, HG004, HG006, and HG007 (5/266); HG00514, HG00731, HG00732, NA19240 from HGSVC2 (Ebert et al. 2021) (4/266); CHM13 (1/266) (Nurk et al. 2022), (https://www.ncbi.nlm.nih.gov/sra/?term=SRX789768*+CHM13)) the HPRC Year 1 and HPRC_PLUS Year 2 releases (101/266)(Ebert et al. 2021). Pbsv calls were merged naively using bcftools merge (v. 1.15.1), i.e., merging only variants that were identical in terms of location, reference sequence, and alternate sequence. The internal pbsv allele frequency callset included affected probands as well as parent/child trios.

A more robustly merged cohort callset (n=96) was constructed using Jasmine, which allows for merging of similar structural variants that may have non-identical representation in terms of genomic position or variant sequence via spanning a structural variant proximity graph. Jasmine was run with options --centroid_merging --min_overlap=0.65 --min_sequence_id=0.75 -- output_genotypes to create the merged callset. This cohort callset was then annotated with five sets of structural variant frequency annotation. Cohort allele frequencies were generated from the merged set with bcftools +fill-tags (set 1, n=96). A second Jasmine merge was used to combine the cohort callset allele count with additional allele frequency resources: the HudsonAlpha internal pbsv callset (described above) (set 2, n=266); gnomAD structural variants (Collins et al. 2020, v4.0, https://gnomad.broadinstitute.org/news/2023-11-v4-structural-variants) (set 3, n=63,046); HGSVC2 structural variants (Ebert et al. 2021) https://ftp.1000genomes.ebi.ac.uk/vol1/ftp/data_collections/HGSVC2/release/v2.0/integrated_callset/variants_freeze4_sv_insdel_alt.vcf.gz) (set 4, n=18); pbsv calls for individuals from the Human Pangenome Reference Consortium and Genome in a Bottle (PacBio; https://github.com/PacificBiosciences/svpack/raw/main/resources/HPRC_GIAB.GRCh38.pbsv.vcf.gz) (set 5, n=103). The second Jasmine merge was run with the prior settings but without the --output_genotypes option and records without genotypes were discarded. A custom python script was used to transfer allele frequency annotations from the annotation source VCFs to the merged VCF based on unique variant identifiers (which were created for each allele frequency resource as needed) and the IDLIST record from Jasmine. The annotated callset was split into individual VCFs with bcftools (bcftools view -I -s <ID>) which were filtered with bcftools to generate the counts in Supplemental Table S2. The “rare” filter described in Supplemental Table S3 was defined as excluding variants with any of the following: within-cohort allele count > 3, in-house allele count > 3, gnomAD maximum population allele frequency (POPMAX_AF) >= 1%, HPRC_GIAB allele frequency >=1%, or HGSVC2 allele frequency >= 1%. The “proband-exclusive” filter described in Supplemental Table S3 was defined as excluding variants with any of the following: within-cohort allele count > 1, in-house allele count > 2, gnomAD allele count > 0, HPRC_GIAB allele count > 0, or HGSVC2 allele count > 0. The in-house allele count cutoff was set at 2 in order to account for the fact that some parental samples were included in the in-house frequency database. Exon regions were defined as RefSeq exons +/-50bp (calculated from bedtools slop). Variant filtering and counting was performed with bcftools view and command-line tools, e.g., bcftools view -R refseq_exons_plus50bp.bed.gz -e ’AC>3 | inhouse_pbsv_AC >3 | gnomad4_POPMAX_AF >= 0.01 | hgsvc2_AF >= 0.01 | hprc_giab_pbsv_AF >= 0.01’ -H <INPUT_FILE> | wc -l for the rare filter and bcftools view -R refseq_exons_plus50bp.bed.gz -e ’AC>1 | inhouse_pbsv_AC>2 | gnomad4_AC>0 | hgsvc2_AC>0 | hprc_giab_pbsv_AC>0’ -H <INPUT_FILE> | wc -l for the exclusive filter.

### Structural Variant Annotation and Curation

For individual case structural variant analysis, a frequency-filtered subset of the proband’s pbsv calls was generated using bcftools annotate and bcftools filter, requiring that calls be located within +/-50bp of a RefSeq exon and have an allele count of < 4 in the HudsonAlpha internal allele frequency dataset (described above). Each call was visualized using a custom pipeline to automatically generate IGV screenshots (Robinson et al. 2011). Additionally, these filtered pbsv variants were annotated, prioritized, and visualized with SvAnna (v1.0.4, annotations v.2204 or v.230, Danis et al. 2022) based on manually curated HPO terms for each case.

### Variant interpretation and orthogonal confirmation

Variant interpretation was performed using ACMG and ClinGen (Richards et al. 2015; Riggs et al. 2020). Variants of interest were either clinically confirmed by the HudsonAlpha Clinical Services Lab, were confirmed within a research lab, and/or were supported by short-read orthogonal data, except where noted in the text (Table 2).

### Sequencing Metrics

Sequencing metrics were generated from the aligned BAMs using the Sentieon implementations of Picard sequencing metrics (Kendig et al. 2019). Sentieon algorithms QualityYield, GCBias, AlignmentStat were run with default settings. Sentieon WgsMetricsAlgo was run with settings -- include_unpaired true true --min_map_qual 0 --min_base_qual 0 --coverage_cap 5000. Further coverage metrics were generated with cramino using the --phased option. Supplemental Table S5 maps each tool or command and output field name to the corresponding entry in Supplemental Tables S1 and S2. SNV and indel counts and ratios were calculated with rtg vcfstats (https://github.com/RealTimeGenomics/rtg-tools). Summary statistics and graphs were calculated with R (v4.3.1), RStudio (v2023.9.1 build 494), and ggplot2 (v3.4.3). Coverage across regions of interest, such as *SHANK3* was calculated using samtools bedcov with default parameters.

### Repeat region detection and analysis

We curated a BED file of disease-associated low-complexity repeat regions in 66 genes from previous studies (Hiatt et al. 2021; Cohen et al. 2022 and references therein). Variant calls from pbsv were extracted from these regions +/- 30bp (Supplemental Table S4). Reads were also visualized using the Integrated Genomics Viewer (IGV). Coverage across low complexity repeat regions was calculated using samtools bedcov with default parameters. A coverage of at least 8x across the low-complexity region was required for inclusion in Supplemental Table S4 and Supplemental Figure S6. We also used TRGT (Dolzhenko et al. 2024) and companion tool TRVZ for visualization of a subset of calls including those for display in Figure S3. TRGT was fed an hg38 reference genome FASTA, BED catalog of tandem repeats, and a sample’s BAM to generate a VCF containing genotypes for each tandem repeat from the provided catalog in the given sample and a BAM containing only reads that span the repeat sequences. Output VCFs and BAMs were sorted, indexed, and fed into TRVZ with the same hg38 reference FASTA and BED catalog of tandem repeats to generate pileup plots for any desired variants.

### 3’ mRNA-seq

Total RNA was isolated from blood samples in PAXgene RNA tubes (PreAnalytiX #762165) according to the manufacturer’s instructions and stored short-term at -20°C. RNA was isolated using the PAX gene Blood RNA Kit (Qiagen #762164) according to the manufacturer’s instructions. Isolated RNA was quantified by the Qubit RNA HS Assay Kit (Thermo Q32855). 425 ng of RNA was used as input for the QuantSeq 3’ mRNA-Seq Library Prep Kit FWD for Illumina and UMI Second Strand Synthesis Module for QuantSeq FWD (Illumina, Read 1) from Lexogen (015.96 and 081.96, respectively). Libraries were quantified using the Qubit DNA HS Assay Kit (Thermo Q32854) and visualized with the BioAnalyzer High Sensitivity DNA Analysis kit (Agilent 5067–4626) and 2100 BioAnalyzer Instrument (Agilent). Sequencing was carried out using Illumina NextSeq 75 bp single-end. UMIs were first extracted from the reads with UMI-tools extract with regex. Reads were then trimmed with bbduk and aligned to hg38-GENCODEv42 using STAR with the Lexogen recommended parameters for QuantSeq. Bams were deduplicated by UMI and mapping coordinates using UMI-tools dedup. Count tables were generated using htseq-count with the intersection-nonempty method.

## Supporting information

Supplemental Case Reports

## Data Availability

For participants who consented to controlled-access sharing, the lrGS data generated in this study will be submitted to dbGAP and/orAnVIL under accession number phs003537.v1 (https://www.ncbi.nlm.nih.gov/projects/gap/cgi-bin/study.cgi?study_id=phs003537.v1).
srGS data for participants who consented to controlled-access sharing in NIH-funded studies are available via dbGAP and/or AnVIL (CSER1: https://www.ncbi.nlm.nih.gov/projects/gap/cgi-bin/study.cgi?study_id=phs001089.v3.p1, dbGaP accession phs001089; SouthSeq: https://anvilproject.org/data/studies/phs002307, dbGaP accession phs002307).

https://www.ncbi.nlm.nih.gov/projects/gap/cgi-bin/study.cgi?study_id=phs001089.v3.p1

https://anvilproject.org/data/studies/phs002307

## DATA ACCESS

For participants who consented to controlled-access sharing, the lrGS data generated in this study will be submitted to dbGAP and/orAnVIL under accession number phs003537.v1 (https://www.ncbi.nlm.nih.gov/projects/gap/cgi-bin/study.cgi?study_id=phs003537.v1). srGS data for participants who consented to controlled-access sharing in NIH-funded studies are available via dbGAP and/or AnVIL (CSER1: https://www.ncbi.nlm.nih.gov/projects/gap/cgi-bin/study.cgi?study_id=phs001089.v3.p1, dbGaP accession phs001089; SouthSeq: https://anvilproject.org/data/studies/phs002307, dbGaP accession phs002307).

## ACKNOWLEDGMENTS

We thank the families who have contributed to our studies and our collaborating physicians and clinical staff for recruitment and enrollment for these research studies. Some reagents were provided by PacBio as part of an early-access testing program. The CSER1 project was supported by a grant from the US National Human Genome Research Institute (NHGRI; UM1HG007301). The SouthSeq project (U01HG007301) was supported by the Clinical Sequencing Evidence-Generating Research (CSER2) consortium, which is funded by the National Human Genome Research Institute with co-funding from the National Institute on Minority Health and Health Disparities and the National Cancer Institute. The Alabama Genomic Health Initiative is an Alabama-State earmarked project (F170303004) through the University of Alabama in Birmingham. The PGEN cohort was funded by the Alabama Pediatric Genomics Initiative. lrGS of some samples was supported by a Research Grant from the Muscular Dystrophy Association (MDA 963255).

