## Supplemental Figures for "Long-read genome sequencing and variant reanalysis increase diagnostic yield in neurodevelopmental disorders"

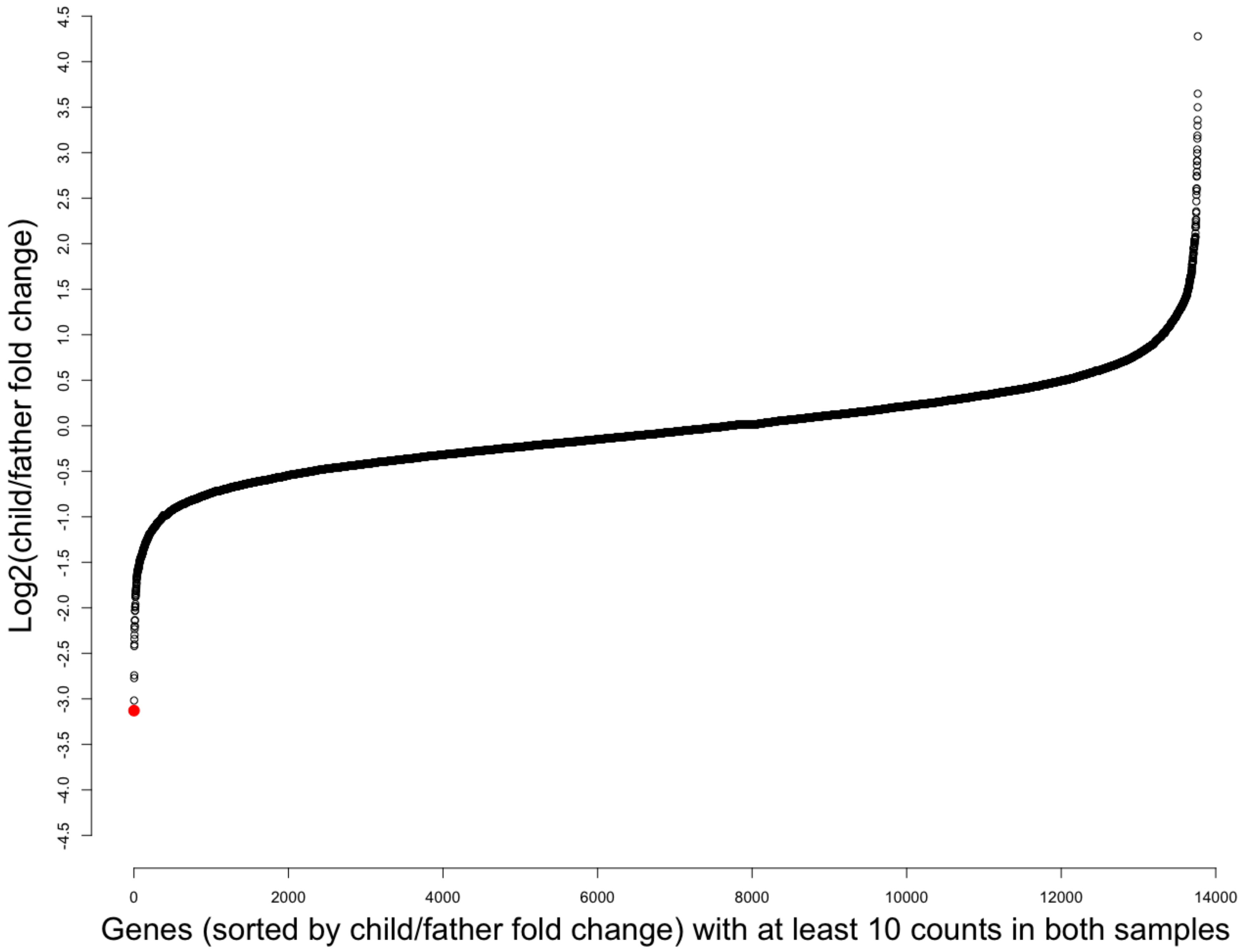


**Supplemental Figure S1. Sorted, relative expression of transcripts in child and father using 3'-end RNA-seq.** *HCFC1* (red dot) shows the greatest expression decrease in the proband relative to his father, an ~8.8-fold reduction. Ten counts in each sample were required.

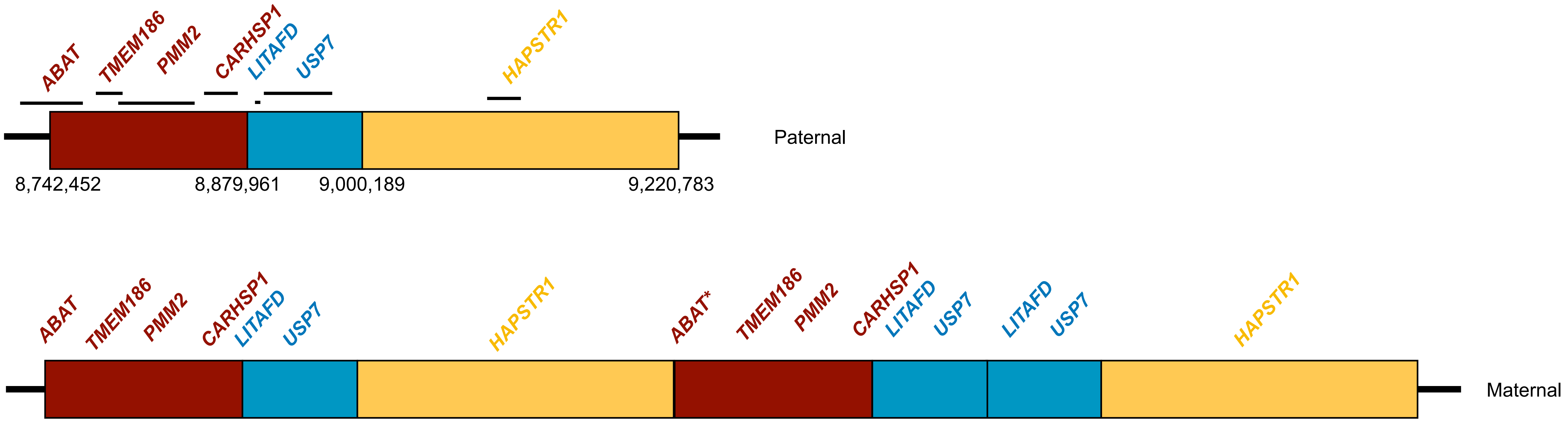


**Supplemental Figure S2. chr16 dup schematic in Proband 4.** Multiple regions are duplicated, resulting in an overall copy number of three for *TMEM186, PMM2, CARHSP1* and *HAPSTR*. Total copy numbers for *LITAFD* and *USP7* are four. Note that the additional copy of ABAT (asterisk) is partial.

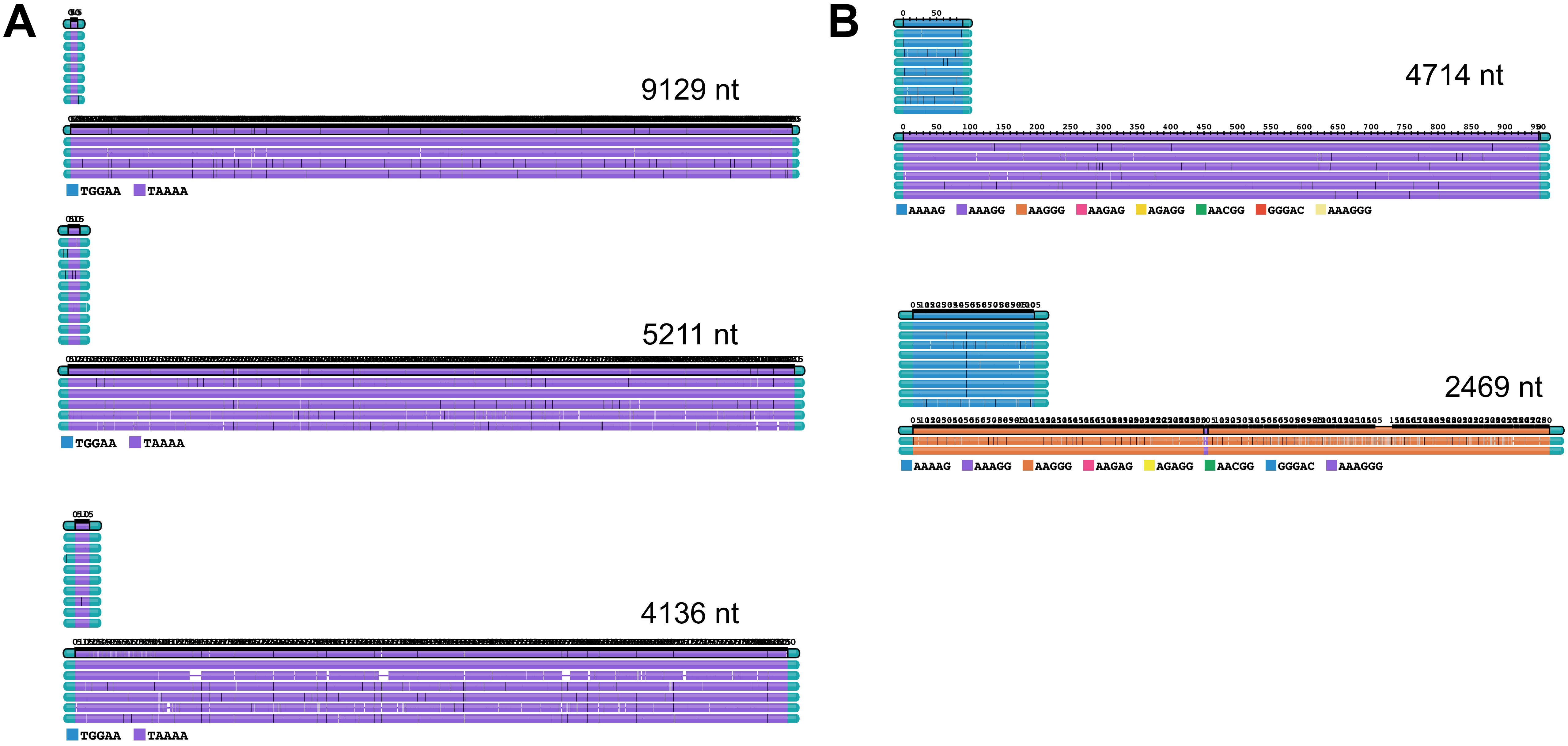


**Supplemental Figure S3. A.** Several large heterozygous insertions in *BEAN1* that were determined to be benign based on sequence content (TAAAA). **B.** Heterozygous expansions in *RFC1* that were determined to be benign (AAAGG) and potentially clinically significant (AAGGG). Note though that only biallelic expansions in *RFC1* have been reported to be associated with disease, and further, this AAGGG repeat is interrupted by a small stretch of AAAGG. Visualization here used TRGT and TRVZ (see Methods).

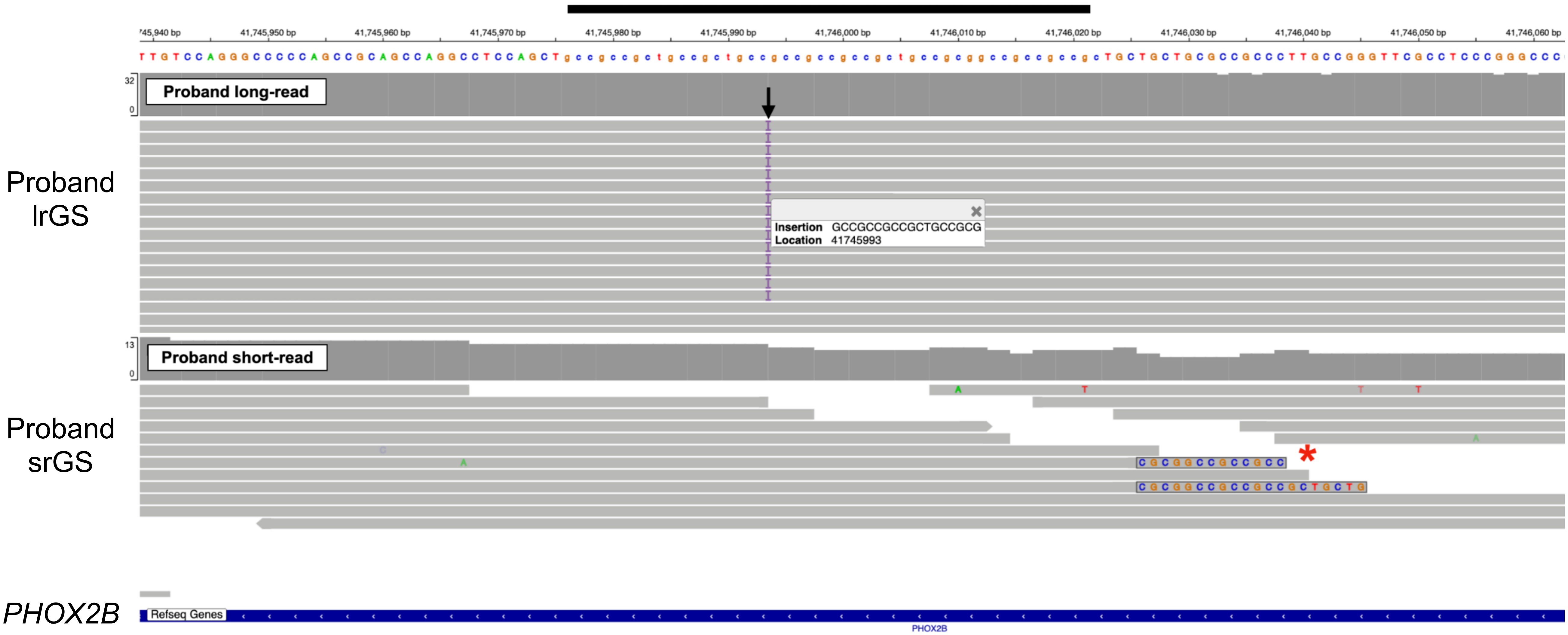


**Supplemental Figure S4.** A pathogenic 18 bp alanine tract expansion in *PHOX2B* (black arrow) was observed in a repeat region (black bar) in Proband 5. Some srGS reads have evidence of this insertion (red asterisk) but the variant was not called.

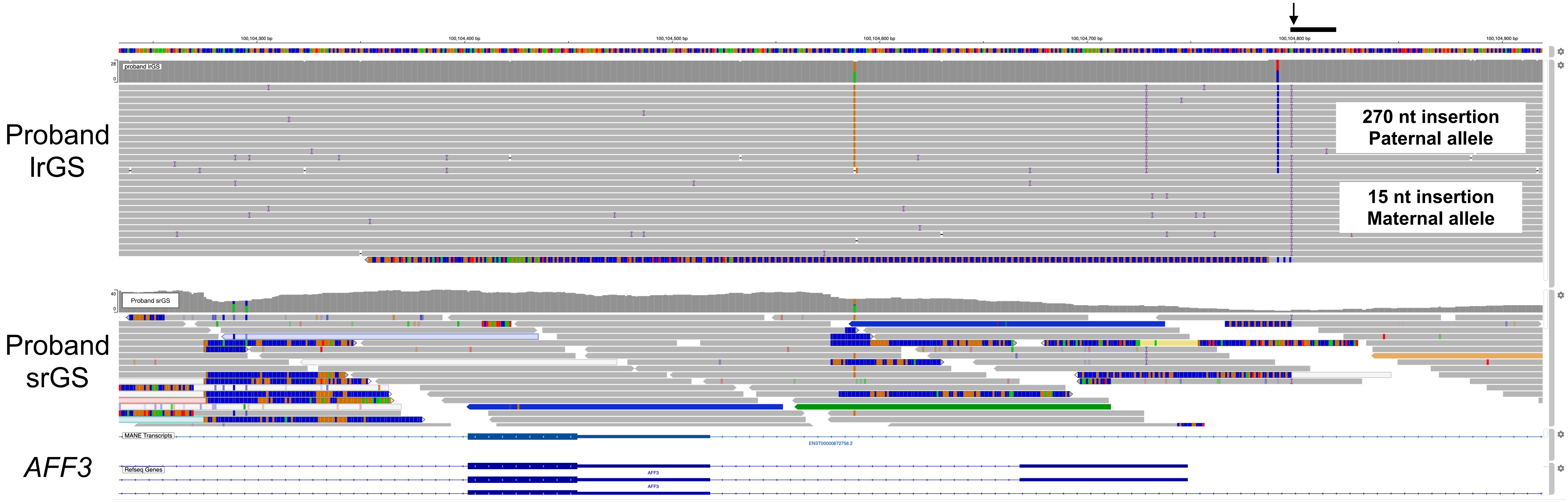


**Supplemental Figure S5.** A 270 bp insertion in the promoter of *AFF3* (black arrow) was observed in a repeat region (black bar) in Proband 6. Segregation of a downstream SNP determined that the 270 bp insertion is on the paternal allele.

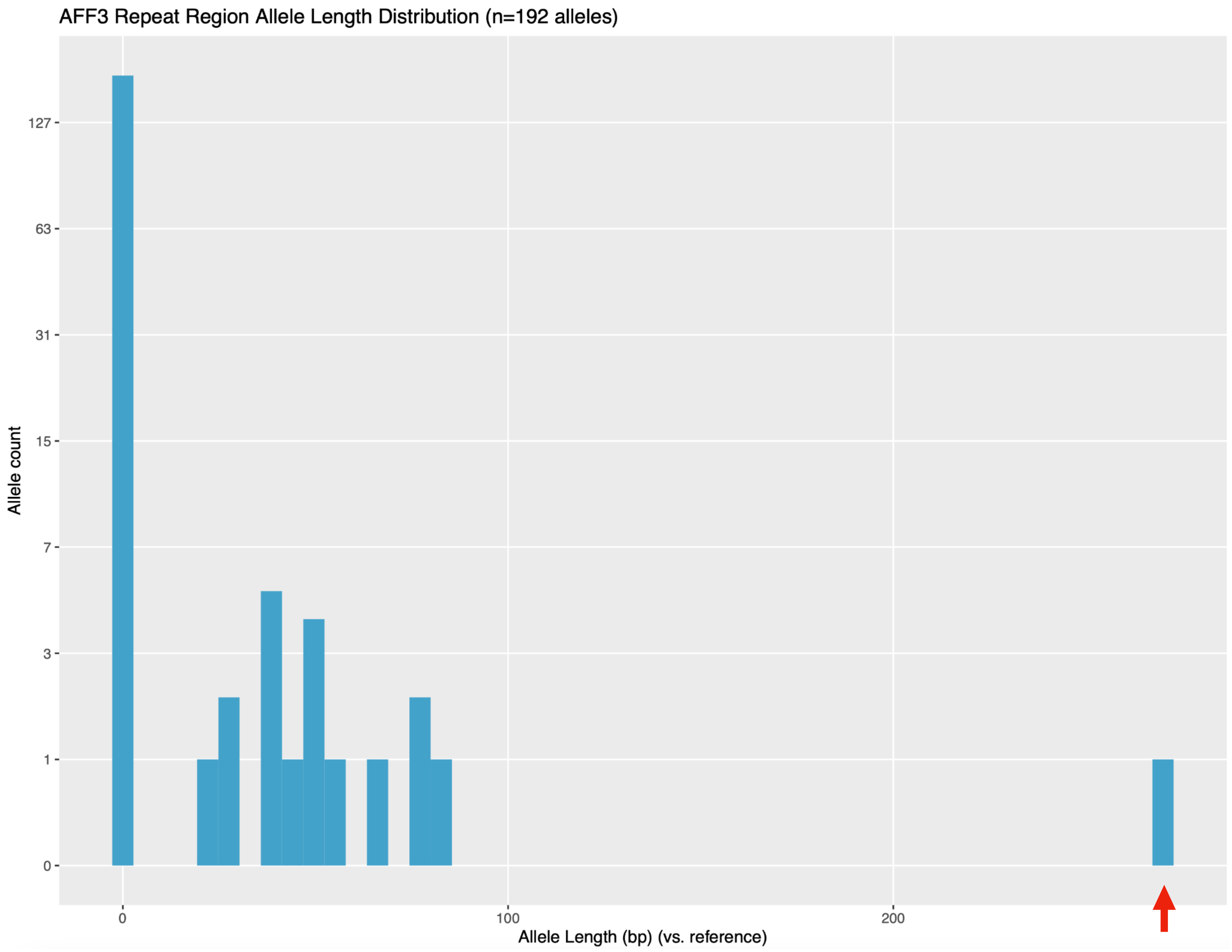


**Supplemental Figure S6.** Distribution of lengths of variant calls in the promoter of *AFF3* (chr2:100104769-100104854) in 96 probands. The 270 nt insertion in *AFF3* in Proband 6 (red arrow) is a clear outlier compared to other calls in long-read data in the region.

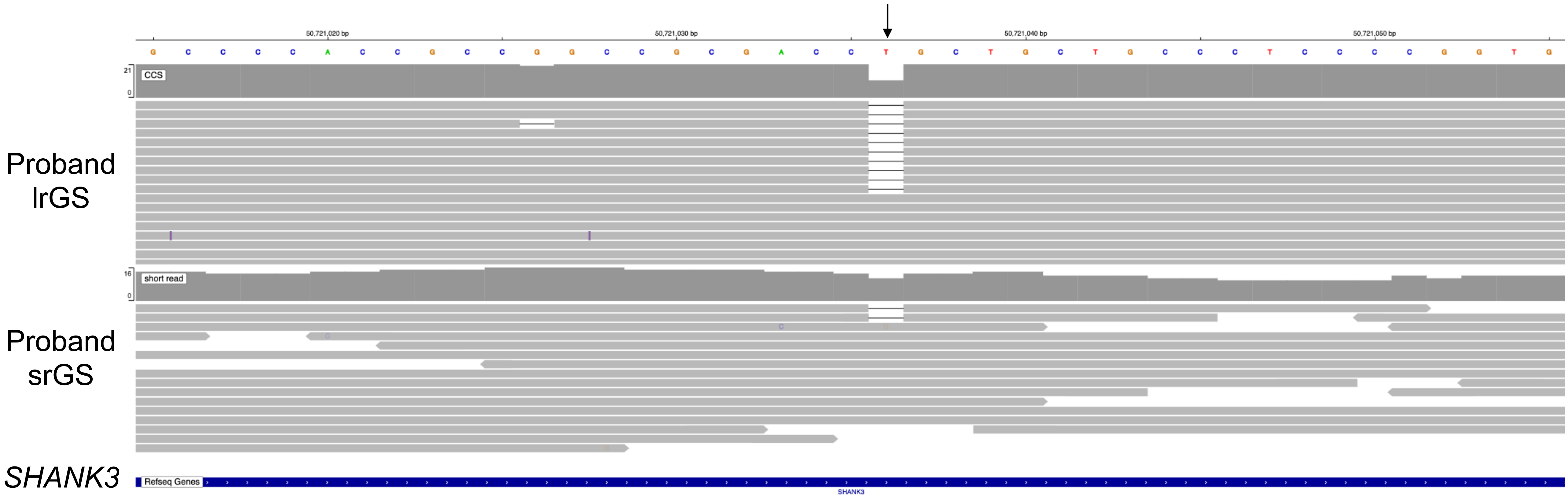


**Supplemental Figure S7.** A 1 bp deletion in *SHANK3* (black arrow) was observed in lrGS data and srGS data but was not called in srGS in Proband 7.

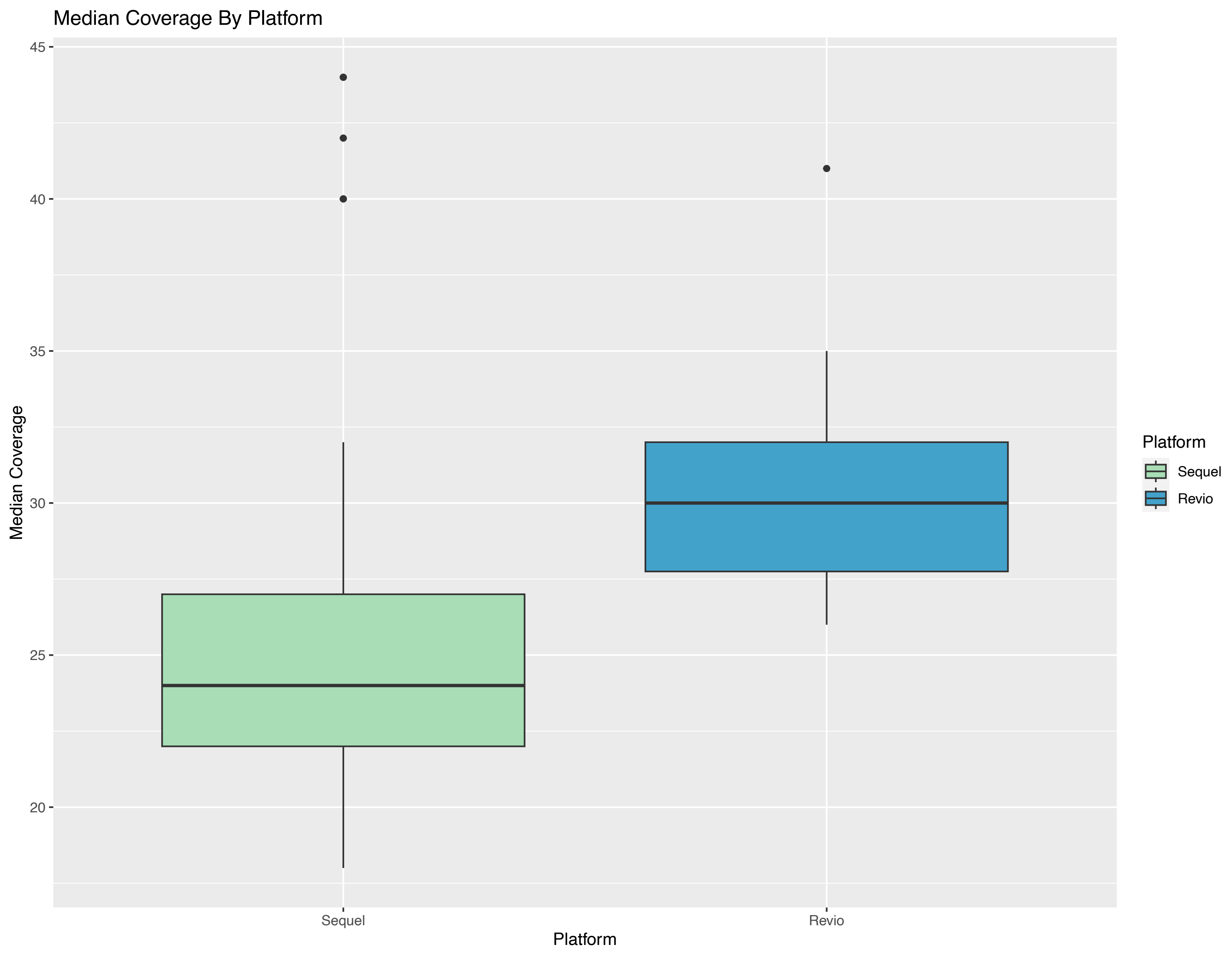


**Supplemental Figure S8.** Median coverage by platform.
