## Supplemental Tables for "Long-read genome sequencing and variant reanalysis increase diagnostic yield in neurodevelopmental disorders"

**Supplemental Table S1. Proband Variant Counts and Sequencing Metrics (n=96)**

| Metric | Mean | Median | Min | Max |
|---|---|---|---|---|
| **Sequencing Metrics (* post/alignment)** | | | | |
| **Sequenced Bases (Gb)** | 82.7 | 80.9 | 53.5 | 131.5 |
| **Read Count** | 5,072,597 | 4,731,572 | 3,008,914 | 8,782,703 |
| **% Q20 Bases** | 97% | 99% | 93% | 99% |
| **% Q30 Bases** | 94% | 97% | 85% | 98% |
| **Mean Read Length (Sequenced)** | 16,485 | 16,771 | 11,295 | 21,412 |
| **hg38 Alignment Metrics** | | | | |
| **Mean Coverage** | 27.8 | 27.3 | 18.1 | 44.2 |
| **Median Coverage** | 27.4 | 27 | 18 | 44 |
| **Percent Covered at 1x** | 99.30% | 99.60% | 98.60% | 99.70% |
| **Percent Covered at 10x** | 97.60% | 97.80% | 95.10% | 99.00% |
| **Percent Covered at 20x** | 80.80% | 84.90% | 36.10% | 97.50% |
| **Percent Covered at 30x** | 35.80% | 33.10% | 1.60% | 92.90% |
| **Aligned Mean Read Length** | 15,648 | 15,916 | 10,955 | 20,100 |
| **Read Phasing Metrics** | | | | |
| **% Reads Phased (WhatsHap)** | 77.20% | 75.80% | 70.50% | 86.90% |
| **Median Phase Block Length** | 282,567 | 131,931 | 14,524 | 3,440,858 |
| **N50 Phase Block Length** | 1,096,824 | 674,583 | 37,430 | 6,163,785 |
| **Small Variant Metrics (DeepVariant)** | | | | |
| **Total Small Variants** | 5,407,963 | 5,375,211 | 4,928,999 | 6,453,286 |
| **SNVs** | 4,413,217 | 4,404,564 | 3,997,350 | 5,299,670 |
| **Total InDels** | 994,746 | 970,031 | 926,286 | 1,153,616 |
| **Insertions** | 487,206 | 475,227 | 445,212 | 572,169 |
| **Deletions** | 482,005 | 465,574 | 445,427 | 568,143 |
| **Complex Indels** | 25,535 | 23,516 | 20,776 | 36,408 |
| **SNV Ti/Tv Ratio** | 1.867 | 1.875 | 1.75 | 1.99 |
| **Het/Hom Ratio** | 1.638 | 1.54 | 1.41 | 2.37 |
| **SNV Het/Hom Ratio** | 1.56 | 1.47 | 1.34 | 2.25 |
| **Insertion Het/Hom Ratio** | 1.98 | 1.85 | 1.74 | 2.82 |
| **Deletion Het/Hom Ratio** | 1.961 | 1.805 | 1.66 | 2.97 |
| **Structural Variant Metrics (pbsv)** | | | | |
| **Total Structural Variant Calls** | 57,203.3 | 55,586.0 | 53,834.0 | 65,120.0 |
| **Deletion** | 25,498.4 | 24,757.0 | 23,799.0 | 29,531.0 |
| **Duplication** | 3,093.8 | 3,016.5 | 2,820.0 | 3,649.0 |
| **Insertion** | 28,275.5 | 27,594.0 | 26,441.0 | 32,437.0 |
| **Inversion** | 121.2 | 121.0 | 99.0 | 155.0 |
| **Breakend** | 200.8 | 193.0 | 124.0 | 312.0 |
| **cnv** | 13.6 | 6.5 | 0.0 | 53.0 |

**Supplemental Table S2. Parent Variant Counts and Sequencing Metrics (n=20)**

| Metric | Mean | Median | Min | Max |
|---|---|---|---|---|
| **Sequencing Metrics (post/alignment)** | | | | |
| **Sequenced Bases (Gb)** | 67.64 | 66.94 | 44.39 | 88.67 |
| **Read Count** | 4,321,913 | 4,232,886 | 2,904,689 | 5,974,978 |
| **% Q20 Bases** | 98.5% | 98.6% | 98.1% | 98.8% |
| **% Q30 Bases** | 96.6% | 96.7% | 95.6% | 97.1% |
| **Mean Read Length (Sequenced)** | 15,805.5 | 16,430.0 | 12,961.0 | 18,408.0 |
| **Mean Coverage** | 22.8 | 22.6 | 15.0 | 29.9 |
| **Median Coverage** | 22.4 | 22 | 15 | 30 |
| **Percent Covered at 1x** | 99.2% | 99.2% | 98.8% | 99.6% |
| **Percent Covered at 10x** | 96.2% | 97.0% | 86.1% | 97.8% |
| **Percent Covered at 20x** | 65.6% | 67.5% | 15.4% | 91.5% |
| **Percent Covered at 30x** | 14.3% | 10.0% | 0.8% | 50.8% |
| **Aligned Mean Read Length** | 15,147 | 15,706 | 12,508 | 17,479 |
| **Read Phasing Metrics (n=18)*** | | | | |
| **% Reads Phased (WhatsHap)** | 77.9% | 77.1% | 72.1% | 86.4% |
| **Median Phase Block Length** | 298,971 | 117,710 | 36,645 | 1,257,823 |
| **N50 Phase Block Length** | 909,429 | 601,919 | 91,195 | 3,624,761 |
| **Small Variant Metrics (DeepVariant)** | | | | |
| **Total Small Variants** | 5,151,077 | 5,010,202 | 4,930,296 | 5,975,303 |
| **SNVs** | 4,181,187 | 4,063,593 | 3,997,600 | 4,855,287 |
| **Total InDels** | 969,890 | 945,131 | 914,206 | 1,120,016 |
| **Insertions** | 472,690 | 461,211 | 447,360 | 541,876 |
| **Deletions** | 473,033 | 463,374 | 432,171 | 544,408 |
| **Complex Indels** | 24,168 | 22,548 | 20,198 | 33,732 |
| **SNV Ti/Tv Ratio** | 1.96 | 1.96 | 1.95 | 1.97 |
| **Het/Hom Ratio** | 1.70 | 1.63 | 1.51 | 2.37 |
| **SNV Het/Hom Ratio** | 1.63 | 1.56 | 1.45 | 2.25 |
| **Insertion Het/Hom Ratio** | 2.00 | 1.92 | 1.78 | 2.79 |
| **Deletion Het/Hom Ratio** | 2.01 | 1.90 | 1.75 | 2.92 |
| **Structural Variant Metrics (pbsv)** | | | | |
| **Total Structural Variants** | 57,650.0 | 56,199.0 | 55,146.0 | 64,546.0 |
| **Deletion** | 25,406.7 | 24,789.0 | 24,302.0 | 29,116.0 |
| **Duplication** | 3,369.0 | 3,294.0 | 3,147.0 | 3,779.0 |
| **Insertion** | 29,032.3 | 28,358.5 | 27,718.0 | 32,234.0 |
| **Inversion** | 355.5 | 353.0 | 335.0 | 392.0 |
| **Breakend** | 390.6 | 390.0 | 346.0 | 434.0 |

*** n=18 on read phasing metrics due to whatshap tags that could not properly be interpreted by cramino to generate bam phasing stats for two samples**

**Supplemental Table S3. Proband Structural Variant Counts (n=96)**

| | All Genomic Regions | | | | Refseq Exons +/- 50bp | | | |
|---|---|---|---|---|---|---|---|---|
| | Mean | Median | Min | Max | Mean | Median | Min | Max |
| **No Allele Frequency Filter** | **56,151.8** | **54,534.5** | **52,990.0** | **64,057.0** | **2,364.7** | **2,294.0** | **2,190.0** | **2,772.0** |
| **Deletions** | 25,019.7 | 24,266.0 | 23,417.0 | 29,022.0 | 1,074.6 | 1,037.0 | 959.0 | 1,278.0 |
| **Duplications** | 3,031.0 | 2,955.5 | 2,765.0 | 3,588.0 | 221.0 | 217.0 | 188.0 | 273.0 |
| **Insertions** | 27,852.2 | 27,170.5 | 25,997.0 | 32,035.0 | 1,038.0 | 1,008.0 | 947.0 | 1,228.0 |
| **Inversions** | 116.9 | 116.0 | 94.0 | 151.0 | 26.7 | 27.0 | 19.0 | 34.0 |
| **Breakends** | 132.0 | 125.5 | 75.0 | 214.0 | 4.4 | 4.0 | 1.0 | 9.0 |
| **Rare (<1% in public SV databases & AC<4 cohort/internal)** | **2,344.7** | **1,720.5** | **1,259.0** | **5,416.0** | **116.6** | **87.0** | **52.0** | **281.0** |
| **Deletions** | 1,297.3 | 933.0 | 679.0 | 3,024.0 | 69.1 | 51.0 | 30.0 | 174.0 |
| **Duplications** | 204.1 | 146.0 | 100.0 | 491.0 | 14.3 | 11.5 | 3.0 | 38.0 |
| **Insertions** | 810.2 | 600.5 | 395.0 | 1,991.0 | 32.1 | 26.0 | 11.0 | 79.0 |
| **Inversions** | 4.1 | 3.0 | 0.0 | 18.0 | 0.6 | 0.0 | 0.0 | 4.0 |
| **Breakends** | 29.1 | 26.5 | 9.0 | 78.0 | 0.5 | 0.0 | 0.0 | 3.0 |
| **Exclusive (Not in public SV databases, cohort/internal AC=1)** | **971.5** | **733.0** | **477.0** | **2,212.0** | **51.1** | **39.5** | **19.0** | **128.0** |
| **Deletions** | 569.5 | 420.0 | 297.0 | 1,279.0 | 31.8 | 25.0 | 12.0 | 80.0 |
| **Duplications** | 80.9 | 59.0 | 32.0 | 203.0 | 5.4 | 4.0 | 0.0 | 18.0 |
| **Insertions** | 302.1 | 231.0 | 116.0 | 780.0 | 13.2 | 11.0 | 2.0 | 34.0 |
| **Inversions** | 2.3 | 1.0 | 0.0 | 13.0 | 0.4 | 0.0 | 0.0 | 4.0 |
| **Breakends** | 16.7 | 15.0 | 4.0 | 56.0 | 0.3 | 0.0 | 0.0 | 3.0 |

**Public DBs: gnomad v4 (n=63,046), HPRC GIAB pbsv calls (PacBio, n=103), HGSVC2 (n=45),**

**These stats are computed per sample from the Jasmine-merged sv callset, so the no allele frequency / all regions values differ slightly from counts reported in Supplemental Table S1.**

**Supplemental Table S4. Repeat region loci (hg38) and allele calls from pbsv in those regions (vs. reference).**

| Chromosome | Repeat Start | Repeat End | Gene | Repeat | Repeat Length | Individuals with coverage >=8 | Min Variant Length (bp) | Median Variant Length (bp) | Mean Variant Length (bp) | 90%ile Length (bp) | 95%ile Length (bp) | 99%ile Length (bp) | Max Allele Length (bp) |
|---|---|---|---|---|---|---|---|---|---|---|---|---|---|
| **chrX** | 148500631 | 148500691 | AFF2 | GCC | 3 | 92 | -24 | 0 | 3 | 0 | 24 | 51 | 81 |
| **chr2** | 100104799 | 100104824 | AFF3 | GCC | 3 | 96 | 0 | 0 | 6 | 0 | 46 | 79 | 270 |
| **chrX** | 67545316 | 67545385 | AR | GCA | 3 | 86 | -24 | 0 | 0 | 0 | 0 | 22 | 24 |
| **chrX** | 25013649 | 25013698 | ARX | GCC | 3 | 91 | 0 | 0 | 0 | 0 | 0 | 0 | 0 |
| **chr12** | 6936716 | 6936773 | ATN1 | CAG | 3 | 96 | -30 | 0 | -2 | 0 | 0 | 0 | 30 |
| **chr6** | 16327633 | 16327723 | ATXN1 | TGC | 3 | 96 | 0 | 0 | 0 | 0 | 0 | 21 | 21 |
| **chr22** | 45795354 | 45795424 | ATXN10 | ATTCT | 5 | 96 | -35 | 0 | 0 | 0 | 0 | 20 | 30 |
| **chr12** | 111598949 | 111599018 | ATXN2 | GCT | 3 | 96 | 0 | 0 | 0 | 0 | 0 | 0 | 21 |
| **chr14** | 92071009 | 92071042 | ATXN3 | CTG | 3 | 96 | 0 | 27 | 22 | 45 | 54 | 64 | 75 |
| **chr3** | 63912684 | 63912726 | ATXN7 | GCA | 3 | 96 | 0 | 0 | 0 | 0 | 0 | 0 | 0 |
| **chr13** | 70139353 | 70139428 | ATXN8 | GCA | 3 | 96 | -21 | 0 | -2 | 0 | 0 | 52 | 156 |
| **chrX** | 19990919 | 19990973 | BCLAF3 | GCC | 3 | 77 | -36 | 0 | 0 | 0 | 0 | 0 | 0 |
| **chr16** | 66490398 | 66490467 | BEAN1 | TGGAA | 5 | 96 | -25 | 20 | 169 | 207 | 467 | 4233 | 9129 |
| **chr11** | 66744821 | 66744850 | C11ORF80 | GCG | 3 | 96 | 0 | 0 | 0 | 0 | 0 | 0 | 0 |
| **chr9** | 27573528 | 27573546 | C9ORF72 | GGCCCC | 6 | 96 | 0 | 0 | 8 | 30 | 42 | 69 | 108 |
| **chr19** | 13207858 | 13207897 | CACNA1A | CAG | 3 | 95 | 0 | 0 | 0 | 0 | 0 | 0 | 0 |
| **chr11** | 119206289 | 119206322 | CBL | CGG | 3 | 96 | 0 | 0 | 0 | 0 | 0 | 0 | 24 |
| **chr3** | 129172576 | 129172732 | CNBP | CAGG | 4 | 96 | -60 | -20 | -14 | 0 | 0 | 0 | 24 |
| **chr19** | 18786034 | 18786050 | COMP | GAC | 3 | 96 | 0 | 0 | 0 | 0 | 0 | 0 | 0 |
| **chr21** | 43776443 | 43776479 | CSTB | CCCCGCC CCGCG | 12 | 96 | 0 | 0 | 2 | 0 | 0 | 0 | 300 |
| **chr12** | 50505001 | 50505022 | DIP2B | CGG | 3 | 96 | 0 | 0 | 8 | 30 | 36 | 61 | 213 |
| **chr19** | 45770204 | 45770264 | DMPK | CAG | 3 | 96 | -45 | -27 | -28 | 0 | 0 | 21 | 27 |
| **chr17** | 80146992 | 80147139 | EIF4A3 | GCCGCTG CCGACCTC GCTGT | 20 | 96 | -58 | 20 | 12 | 40 | 60 | 60 | 102 |
| **chrX** | 147912050 | 147912110 | FMR1 | GGC | 3 | 88 | -30 | 30 | 24 | 36 | 36 | 60 | 69 |
| **chr3** | 138946020 | 138946063 | FOXL2 | GCG | 3 | 96 | 0 | 0 | 0 | 0 | 0 | 0 | 0 |
| **chr10** | 93702522 | 93702547 | FRA10AC1 | CCG | 3 | 96 | 0 | 0 | 3 | 0 | 21 | 30 | 292 |
| **chr9** | 69037261 | 69037304 | FXN | GAA | 3 | 96 | 0 | 0 | 4 | 19 | 41 | 55 | 79 |
| **chr19** | 14496041 | 14496074 | GIPC1 | CGG | 3 | 96 | 0 | 0 | 0 | 0 | 0 | 2 | 24 |
| **chr2** | 190880872 | 190880920 | GLS | GCA | 3 | 96 | -24 | 0 | -7 | 0 | 0 | 2 | 21 |
| **chr7** | 27199825 | 27199862 | HOXA13 | GCC | 3 | 96 | 0 | 0 | 0 | 0 | 0 | 0 | 0 |
| **chr2** | 176093058 | 176093104 | HOXD13 | GGC | 3 | 96 | 0 | 0 | 0 | 0 | 0 | 0 | 0 |
| **chr4** | 3074876 | 3074966 | HTT | CAG | 3 | 96 | -24 | 0 | 1 | 0 | 0 | 36 | 42 |
| **chr16** | 87604287 | 87604329 | JPH3 | GCT | 3 | 96 | 0 | 0 | 0 | 0 | 0 | 27 | 33 |
| **chr4** | 21713786 | 21716097 | KCNIP4 | TAAAA | 3 | 96 | -257 | -237 | -160 | 0 | 0 | 149 | 459 |
| **chr8** | 104588972 | 104589000 | LRP12 | CGG | 3 | 96 | 0 | 0 | 0 | 0 | 0 | 0 | 21 |
| **chr5** | 10356346 | 10356412 | MARCHF6 | TTTCA | 5 | 95 | -20 | 0 | 0 | 0 | 0 | 3 | 30 |
| **chr3** | 151367585 | 151369763 | MED12 | ATTTC | 5 | 96 | -110 | -65 | -49 | 0 | 0 | 0 | 0 |
| **chr9** | 110700652 | 110702927 | MUSK | TTTAT | 5 | 96 | -222 | -94 | -57 | 61 | 139 | 895 | 4262 |
| **chr15** | 22786677 | 22786701 | NOP56 | GGGCCT | 6 | 96 | 0 | 0 | 3 | 24 | 24 | 25 | 30 |
| **chr20** | 2652733 | 2652775 | NOP56 | GGCCTG | 6 | 96 | 0 | 0 | 3 | 24 | 24 | 25 | 30 |

| | | | | | | | | | | | | |
|---|---|---|---|---|---|---|---|---|---|---|---|---|
| **chr1** | 146228800 | 146228821 | NOTCH2NLA | GCC | 3 | 93 | 0 | 0 | 0 | 0 | 0 | 0 | 0 |
| **chr1** | 149390802 | 149390841 | NOTCH2NLC | GGC | 3 | 96 | 0 | 0 | 12 | 27 | 31 | 42 | 51 |
| **chr10** | 79826383 | 79826404 | NUTM2B-AS1 | CGG | 3 | 95 | 0 | 0 | 1 | 0 | 19 | 19 | 24 |
| **chr14** | 23321472 | 23321490 | PABPN1 | GCG | 3 | 96 | 0 | 0 | 0 | 0 | 0 | 0 | 0 |
| **chr4** | 41745972 | 41746032 | PHOX2B | GCC | 3 | 96 | -21 | 0 | 0 | 0 | 0 | 0 | 0 |
| **chr5** | 146878727 | 146878757 | PPP2R2B | GCT | 3 | 96 | 0 | 0 | 1 | 0 | 0 | 28 | 30 |
| **chr9** | 130681606 | 130681641 | PRDM12 | GCN | 3 | 96 | 0 | 0 | 0 | 0 | 0 | 0 | 0 |
| **chr20** | 4699379 | 4699380 | PRNP | GCAGCCT CATGGTG GTGGCTG GGGGCAG CCCCAT | 34 | 96 | 0 | 0 | 0 | 0 | 0 | 0 | 0 |
| **chr4** | 159342526 | 159342617 | RAPGEF2 | TTTCA | 5 | 96 | -20 | 0 | 1 | 0 | 0 | 25 | 40 |
| **chr4** | 39348424 | 39348479 | RFC1 | AAAAG | 5 | 92 | -20 | 0 | 187 | 502 | 554 | 1043 | 4714 |
| **chr6** | 45422750 | 45422802 | RUNX2 | GGC | 3 | 96 | 0 | 0 | 0 | 0 | 0 | 0 | 0 |
| **chr8** | 118366815 | 118366919 | SAMD12 | AAATA | 5 | 96 | -49 | 0 | -5 | 0 | 0 | 2 | 20 |
| **chrX** | 140504316 | 140504362 | SOX3 | GGC | 3 | 94 | 0 | 0 | 0 | 0 | 0 | 0 | 0 |
| **chr2** | 96197066 | 96197122 | STARD7 | ATTTC | 5 | 96 | 0 | 0 | 18 | 25 | 82 | 354 | 867 |
| **chr6** | 170561906 | 170562017 | TBP | GCA | 3 | 96 | -21 | 0 | 0 | 0 | 0 | 0 | 0 |
| **chr18** | 55586155 | 55586227 | TCF4 | TGC | 3 | 96 | -39 | -24 | -10 | 0 | 96 | 178 | 225 |
| **chrX** | 149631763 | 149631782 | TMEM185A | GCC | 3 | 92 | -33 | -21 | -18 | 0 | 0 | 37 | 42 |
| **chr16** | 24613439 | 24613530 | TNRC6A | TTTCA | 5 | 96 | -35 | -25 | -23 | 0 | 0 | 0 | 0 |
| **chr1** | 1435799 | 1435820 | TRIO | ATTTT | 6 | 96 | 0 | 0 | 0 | 0 | 0 | 0 | 20 |
| **chr16** | 17470907 | 17470923 | VWA1 | GGCGCGG AGC | 10 | 77 | 0 | 0 | 0 | 0 | 0 | 0 | 0 |
| **chr3** | 183712187 | 183712223 | XYLT1 | CGG | 3 | 96 | 0 | 238 | 234 | 265 | 271 | 275 | 280 |
| **chr16** | 72787694 | 72787757 | YEATS2 | TTTTA | 5 | 96 | 0 | 0 | 28 | 153 | 153 | 158 | 163 |
| **chr13** | 99985448 | 99985494 | ZDHHC14 | ATTTC | 5 | 96 | -35 | 0 | -5 | 0 | 0 | 0 | 0 |
| **chr7** | 55887601 | 55887640 | ZFHX3 | RCC | 3 | 96 | -30 | 0 | 0 | 0 | 0 | 0 | 0 |
| **chr5** | 14420230 | 14422285 | ZIC2 | GCG | 3 | 96 | -27 | 0 | 0 | 0 | 0 | 0 | 0 |
| **chr6** | 157533533 | 157535600 | ZNF713 | CGG | 3 | 95 | -94 | 0 | 0 | 0 | 0 | 35 | 51 |

**Supplemental Table S5. Sequencing Metrics Sources**

<table>
<tr><th>Metric</th><th>Algorithm</th><th>Output FIeld Name</th></tr>
<tr><td>Sequencing Metrics (* post/alignment)</td><td></td><td></td></tr>
<tr><td>Sequenced Bases (Gb)</td><td>sentieon --algo QualityYield</td><td>TOTAL_BASES</td></tr>
<tr><td>Read Count</td><td>sentieon --algo QualityYield</td><td>TOTAL_READS</td></tr>
<tr><td>% Q20 Bases</td><td>sentieon --algo QualityYield</td><td>Q20_BASES</td></tr>
<tr><td>% Q30 Bases</td><td>sentieon --algo QualityYield</td><td>Q30_BASES</td></tr>
<tr><td>Mean Read Length (Sequenced)</td><td>sentieon --algo QualityYield</td><td>READ_LENGTH</td></tr>
<tr><td>Mean Coverage</td><td>sentieon --algo WgsMetricsAlgo</td><td>MEAN_COVERAGE</td></tr>
<tr><td>Median Coverage</td><td>sentieon --algo WgsMetricsAlgo</td><td>MEDIAN_COVERAGE</td></tr>
<tr><td>Percent Covered at 1x</td><td>sentieon --algo WgsMetricsAlgo</td><td>PCT_1X</td></tr>
<tr><td>Percent Covered at 10x</td><td>sentieon --algo WgsMetricsAlgo</td><td>PCT_10X</td></tr>
<tr><td>Percent Covered at 20x</td><td>sentieon --algo WgsMetricsAlgo</td><td>PCT_20X</td></tr>
<tr><td>Percent Covered at 30x</td><td>sentieon --algo WgsMetricsAlgo</td><td>PCT_30X</td></tr>
<tr><td>Aligned Mean Read Length</td><td>cramino</td><td>Mean Length</td></tr>
<tr><td>Read Phasing Metrics</td><td></td><td></td></tr>
<tr><td>% Reads Phased (WhatsHap)</td><td>cramino</td><td>Fraction reads phased</td></tr>
<tr><td>Median Phase Block Length</td><td>cramino</td><td>Median phaseblock length</td></tr>
<tr><td>N50 Phase Block Length</td><td></td><td>N50 Phaseblock Length</td></tr>
<tr><td>Small Variant Metrics (DeepVariant)</td><td></td><td></td></tr>
<tr><td>Total Small Variants</td><td>rtg vcfstats</td><td>Passed_Filters</td></tr>
<tr><td>SNVs</td><td>rtg vcfstats</td><td>SNPs</td></tr>
<tr><td>Total InDels</td><td>rtg vcfstats</td><td>(Passed_Filters - SNPs)</td></tr>
<tr><td>Insertions</td><td>rtg vcfstats</td><td>Insertions</td></tr>
<tr><td>Deletions</td><td>rtg vcfstats</td><td>Deletions</td></tr>
<tr><td>Complex Indels</td><td>rtg vcfstats</td><td>Indels</td></tr>
<tr><td>SNV Ti/Tv Ratio</td><td>rtg vcfstats</td><td>SNP_Transitions_Transversions</td></tr>
<tr><td>Het/Hom Ratio</td><td>rtg vcfstats</td><td>Total_Het_Hom_ratio</td></tr>
<tr><td>SNV Het/Hom Ratio</td><td>rtg vcfstats</td><td>SNP_Het_Hom_ratio</td></tr>
<tr><td>Insertion Het/Hom Ratio</td><td>rtg vcfstats</td><td>Insertion_Het_Hom_ratio</td></tr>
<tr><td>Deletion Het/Hom Ratio</td><td>rtg vcfstats</td><td>Deletion_Het_Hom_ratio</td></tr>
<tr><td>Structural Variant Metrics (pbsv)</td><td></td><td></td></tr>
<tr><td>Total Structural Variant Calls</td><td>bcftools view -H <input.vcf> | wc -l</td><td>N/A</td></tr>
<tr><td>Deletion</td><td>bcftools filter -i 'SVTYPE==DEL' <input.vcf> | wc -l</td><td>N/A</td></tr>
<tr><td>Duplication</td><td>bcftools filter -i 'SVTYPE==DUP' <input.vcf> | wc -l</td><td>N/A</td></tr>
<tr><td>Insertion</td><td>bcftools filter -i 'SVTYPE==INS' <input.vcf> | wc -l</td><td>N/A</td></tr>
<tr><td>Inversion</td><td>bcftools filter -i 'SVTYPE==INV' <input.vcf> | wc -l</td><td>N/A</td></tr>
<tr><td>Breakend</td><td>bcftools filter -i 'SVTYPE==BND' <input.vcf> | wc -l</td><td>N/A</td></tr>
<tr><td>cnv</td><td>bcftools filter -i 'SVTYPE==cnv' <input.vcf> | wc -l</td><td>N/A</td></tr>
<tr><td colspan="3">Note: sentieon metrics are an implementation of Picard metrics defined here: https://broadinstitute.github.io/picard/picard-metric-definitions.html</td></tr>
</table>
