## Supplemental Case Reports for "Long-read genome sequencing and variant reanalysis increase diagnostic yield in neurodevelopmental disorders"

*HNRNPU*

Proband 8 has a *de novo* variant in *HNRNPU* (NM_031844.3:c.660_661dupAGGCGGCGGA, p.(Gly221ArgfsTer25)). Loss-of-function variation in *HNRNPU* has been associated with Developmental and epileptic encephalopathy 54 (DEE54), which was known at the time of first analysis. However, this variant only affects one isoform (NM_031844.2), and at the time it was not considered a canonical transcript. Interestingly, mouse *Hnrnpu* did not have support for an orthologous transcript. In reviewing the lrGS data, we found that since original analysis, this variant has been reported as pathogenic in ClinVar by two other groups (https://www.ncbi.nlm.nih.gov/clinvar/variation/1174635/). Further, the splice pattern is now supported by CCDS41479.1. The proband is reported to have hypotonia, seizures, speech delay, developmental delay, and mild intellectual disability, features that are consistent with DEE54. We now classify this isoform-specific variant as pathogenic, with the case-level designation as Definitive Diagnostic.

*CSNK2B*

Proband 9 has a paternally-inherited variant in *CSNK2B* (NM_001320.6:c.202C>T, p.(Gln68Ter)). The proband first underwent srGS in 2017, and no findings were returned. Later that same year, loss-of-function variation in *CSNK2B* was reported in probands with ID, with or without epilepsy (Poirier et al. 2017). Reanalysis of the proband's lrGS data identified this likely pathogenic variant. The proband's father is reported to have had special education classes. The case-level designation is Likely Diagnostic.

*GNB2*

Proband 10 has a maternally-inherited missense variant in *GNB2* (NM_005273.4:c.217G>A, p.(Ala73Thr)). This individual had singleton srGS in 2021, and no results were returned. Several months after analysis, this variant was submitted to ClinVar as pathogenic by two submitters, and in 2022, a publication suggested this was a recurrent pathogenic variant, reported as a *de novo* variant in three cases (Tan et al. 2022). Reinterpretation of this variant led to classification as likely pathogenic. However, we note that the variant is maternally inherited, as opposed to the *de novo* reports in literature. The proband's mother is not reported to have any overlapping features to reported cases, but Tan, et al. suggest that this variant may lead to milder presentation and mild developmental delays. The proband does have some overlapping features when comparing to the entire cohort (e.g. microcephaly and contractures/hypotonia), but we do not have updates on the development of this young proband. For this reason, we have assigned a case-level designation of Uncertain.

*MCF2*

Proband 11 has a maternally-inherited variant in *MCF2* (NM_005369.5:c.2234G>T, p.(Gly745Val)). At the time of first analysis in 2018, no results were returned. Since that time, a publication identified a proband with a missense variant in *MCF2* ((NM_005369.5): c.4G>A, p.(Ala2Thr), (Molinard-Chenu et al. 2020). The reported proband has congenital bilateral perisylvian syndrome (CBPS) with lower motor neuron dysfunction.  Based on these limited reports, we submitted this variant to GeneMatcher following lrGS. Other cases with *MCF2* missense variants were identified that overlap for developmental delay and hypotonia. We consider this a VUS in a gene of uncertain clinical significance, and we are pursuing additional information. The case-level designation is Uncertain.

*NOTCH3*
Proband 12 harbors a heterozygous, paternally-inherited variant in *NOTCH3* (NM_000435.3c.6409_6410delCT, p.(Leu2137GlyfsTer104)). Variation in *NOTCH3* is associated with several disorders, most notably Lateral Meningocele Syndrome (MIM: 130720), which associates with a variety of muscular, neurological, developmental, and craniofacial symptoms. This variant was not initially flagged in the original analysis because it was inherited from a parent who was not reported to be affected. However, after flagging this variant in the IrGS data and obtaining more family history information, the father indicated that as a child he had congenital lung problems and muscle weakness that resolved in childhood. The variant has been classified as likely pathogenic with a case-level designation of Likely Diagnostic.

*AFF4*
Proband 13 has a paternally-inherited frameshift in *AFF4* (NM_031844.3:c.660_661dupAGGCGGCGGA, p.(Gly221ArgfsTer25)). Missense variants with a presumed gain-of-function effect have been associated with CHOPS Syndrome, named for cognitive impairment, coarse facies, heart defects, obesity, pulmonary involvement, short stature, and skeletal dysplasia (Izumi et al. 2015). This individual has Autism, moderate ID, speech delay, fine motor delays, craniosynostosis, and mild facial dysmorphism. He has had a normal brain MRI. The father has no reported symptoms. While no LOF variants have been reported for this gene, it does appear to be very intolerant to loss-of-function. Following analysis of this variant in IrGS data, we contacted the authors of the paper above, and have submitted this gene to GeneMatcher. We are aware of several other probands with predicted LOF variation, and investigations are ongoing. We have classified this variant as a VUS in a gene of uncertain clinical significance, with a case-level designation of Uncertain.

*KIF1A, KCNT2*
Proband 14 has two SNVs in KIF21A: a paternally-inherited missense (NM_017641.3:c.847C>T, p.(Arg283Cys)) and a maternally-inherited premature stop (NM_017641.3:c.706C>T, p.(Gln236Ter)). While missense variants have previously been associated with congenital fibrosis of extra ocular muscles, biallelic loss of function variation has recently been associated with severe fetal akinesia and arthrogryposis multiplex (Falb et al. 2021). As this proband has a missense and premature stop, it is possible that they present with a milder phenotype. We have submitted this to GeneMatcher and have contacted the authors of the above paper. A collaboration leading to the description of multiple additional individuals with variation in *KIF1A* is underway. We have classified the missense variant as a VUS and the premature stop as likely pathogenic.

This proband also has a paternally-inherited, ~15.3 kb duplication of four exons of *KCNT2* (NC_000001.11:196329420-196344697_DUP). This duplication is predicted to lead to loss-of-function. Variants in *KCNT2* have been associated with Developmental and epileptic encephalopathy 57. Most reported variants are de novo, and most are missense, but LOF variation has been reported as pathogenic in ClinVar. While the gene appears to be relatively intolerant to LOF variation, it is unclear if LOF is truly a pathogenic mechanism of disease for this gene. We have classified this duplication as a VUS.

This proband had trio srGS in 2016, and had no variants of interest returned. More recent publications of both *KCNT2* (Gururaj et al. 2017; Ambrosino et al. 2018) and *KIF21A* (Falb et al. 2021) led to reinterpretation of these variants. Note that the *KIF21A* variants have been clinically validated but the *KCNT2* duplication has not. It is supported by srGS data. The overall case-level designation is Uncertain.

*NRXN1*

Proband 15 has a heterozygous, *de novo* deletion affecting the last exon of *NRXN1* (NC_000002.12:49922063_49928691del). Heterozygous deletions of *NRXN1* have been reported in individuals with autism, schizophrenia, and intellectual disability, although variable phenotypic expressivity and incomplete penetrance have been described (Gauthier et al. 2011; Béna et al. 2013; Walsh et al. 2008; Bucan et al. 2009). During the first analysis of this data, the evidence supporting a highly penetrant NDD from this presumed LOF variant was unclear, However, the accumulation of data since that time makes the return of this variant more compelling (Tromp et al. 2021; Williams et al. 2019). This variant has not been clinically validated but is supported by srGS data. The deletion is classified as a VUS and case-level designation is Uncertain.

*SCN1A*

Proband 16 had srGS in 2017 but no variants of interest were identified. lrGS also revealed no variants of interest, but subsequent reanalysis searching for potential variants affecting poison exon (PE) inclusion revealed a paternally-inherited, deep intronic variant in *SCN1A* (NM_001165963.4:c.4003-603T>C) that may affect splicing of *SCN1A* transcripts (Felker et al. 2023). Pathogenic variants in this gene are associated with multiple disorders, including Dravet syndrome (MIM: 607208). The proband has intellectual disability, developmental delay, and seizures and a strong family history of seizures and suspected Dravet Syndrome. The variant was inherited from the father, who reported a history of seizures. An affected sibling also shares this variant. The proband has an additional deceased sibling who had seizures and whose variant status is unknown. This variant has been classified as a VUS due to lack of functional confirmation at this time. Case-level designation is Uncertain.

### Supplemental Case Reports References

Ambrosino P, Soldovieri MV, Bast T, Turnpenny PD, Uhrig S, Biskup S, Döcker M, Fleck T, Mosca I, Manocchio L, et al. 2018. De novo gain-of-function variants in KCNT2 as a novel cause of developmental and epileptic encephalopathy. *Ann Neurol* **83**: 1198–1204.

Béna F, Bruno DL, Eriksson M, van Ravenswaaij-Arts C, Stark Z, Dijkhuizen T, Gerkes E, Gimelli S, Ganesamoorthy D, Thuresson AC, et al. 2013. Molecular and clinical characterization of 25 individuals with exonic deletions of NRXN1 and comprehensive review of the literature. *American Journal of Medical Genetics, Part B: Neuropsychiatric Genetics* **162**: 388–403.

Bucan M, Abrahams BS, Wang K, Glessner JT, Herman EI, Sonnenblick LI, Alvarez Retuerto AI, Imielinski M, Hadley D, Bradfield JP, et al. 2009. Genome-wide analyses of exonic copy number variants in a family-based study point to novel autism susceptibility genes. *PLoS Genet* **5**.

Falb RJ, Müller AJ, Klein W, Grimmel M, Grasshoff U, Spranger S, Stöbe P, Gauck D, Kuechler A, Dikow N, et al. 2021. Bi-allelic loss-of-function variants in KIF21A cause severe fetal akinesia with arthrogryposis multiplex. *J Med Genet* **60**: 48–56.

Felker SA, Lawlor JMJ, Hiatt SM, Thompson ML, Latner DR, Finnila CR, Bowling KM, Bonnstetter ZT, Bonini KE, Kelly NR, et al. 2023. Poison exon annotations improve the yield of clinically relevant variants in genomic diagnostic testing. *Genet Med* 100884. https://pubmed.ncbi.nlm.nih.gov/37161864/ (Accessed May 29, 2023).

Gauthier J, Siddiqui TJ, Huashan P, Yokomaku D, Hamdan FF, Champagne N, Lapointe M, Spiegelman D, Noreau A, Lafrenière RG, et al. 2011. Truncating mutations in NRXN2 and NRXN1 in autism spectrum disorders and schizophrenia. *Hum Genet* **130**: 563–73. http://www.ncbi.nlm.nih.gov/pubmed/21424692 (Accessed February 15, 2024).

Gururaj S, Palmer EE, Sheehan GD, Kandula T, Macintosh R, Ying K, Morris P, Tao J, Dias KR, Zhu Y, et al. 2017. A De Novo Mutation in the Sodium-Activated Potassium Channel KCNT2 Alters Ion Selectivity and Causes Epileptic Encephalopathy. *Cell Rep* **21**: 926–933.

Izumi K, Nakato R, Zhang Z, Edmondson AC, Noon S, Dulik MC, Rajagopalan R, Venditti CP, Gripp K, Samanich J, et al. 2015. Germline gain-of-function mutations in AFF4 cause a developmental syndrome functionally linking the super elongation complex and cohesin. *Nat Genet* **47**: 338–344. https://pubmed.ncbi.nlm.nih.gov/25730767/ (Accessed February 15, 2024).

Molinard-Chenu A, Fluss J, Laurent S, Laurent M, Guipponi M, Dayer AG. 2020. MCF2 is linked to a complex perisylvian syndrome and affects cortical lamination. *Ann Clin Transl Neurol* **7**: 121–125. https://pubmed.ncbi.nlm.nih.gov/31846234/ (Accessed February 15, 2024).

Poirier K, Hubert L, Viot G, Rio M, Billuart P, Besmond C, Bienvenu T. 2017. CSNK2B splice site mutations in patients cause intellectual disability with or without myoclonic epilepsy. *Hum Mutat* **38**: 932–941. https://pubmed.ncbi.nlm.nih.gov/28585349/ (Accessed February 15, 2024).

Tan NB, Pagnamenta AT, Ferla MP, Gadian J, Chung BHY, Chan MCY, Fung JLF, Cook E, Guter S, Boschann F, et al. 2022. Recurrent de novo missense variants in GNB2 can cause syndromic intellectual disability. *J Med Genet* **59**: 511–516. https://pubmed.ncbi.nlm.nih.gov/34183358/ (Accessed February 15, 2024).

Tromp A, Mowry B, Giacomotto J. 2021. Neurexins in autism and schizophrenia—a review of patient mutations, mouse models and potential future directions. *Mol Psychiatry* **26**: 747–760.

Walsh T, McClellan JM, McCarthy SE, Addington AM, Pierce SB, Cooper GM, Nord AS, Kusenda M, Malhotra D, Bhandari A, et al. 2008. Rare structural variants disrupt multiple genes in neurodevelopmental pathways in schizophrenia. *Science (1979)* **320**: 539–543.

Williams SM, An JY, Edson J, Watts M, Murigneux V, Whitehouse AJO, Jackson CJ, Bellgrove MA, Cristino AS, Claudianos C. 2019. An integrative analysis of non-coding regulatory DNA variations associated with autism spectrum disorder. *Mol Psychiatry* **24**: 1707–1719.
